# Spatial modelling of sand fly vector’s response to a synthetic sex-aggregation pheromone: impact on the incidence of visceral leishmaniasis in rural and urban settings

**DOI:** 10.1101/2020.10.18.20214569

**Authors:** Renata Retkute, Erin Dilger, James GC Hamilton, Matt J Keeling, Orin Courtenay

## Abstract

**Background:** Visceral leishmaniasis (VL) is a potentially fatal protozoan parasitic disease of humans and dogs. In the Americas, dogs are the reservoir and *Lutzomyia longipalpis* is the sand fly vector. A synthetic version of the vector’s sex-aggregation pheromone attracts conspecifics to co-located lethal insecticide, reducing reservoir infection and vector abundance. Mathematical models of spatially deployed interventions are lacking, thus best practise using this novel lure-and-kill vector control approach to reduce infection incidence has not been fully explored.

**Methods:** We developed a predictive mathematical model of vector host-seeking behaviour combined with spatially explicit transmission models to evaluate changes in human and canine spatial infection incidence under variable pheromone implementation scenarios and demographic conditions.

**Results:** The risk of human infection increased exponentially with canine incidence, but at different rates between rural and urban settings with spatial clustering of high forces of human infection related to their proximity to canine infected households. A predicted 70% household coverage using a cost-effective amount (50mg) of pheromone, plus insecticide, reduced rural and urban setting infection incidence by approximately 44% and 50% in dogs, and by 64% and 68% in humans, within 1-2 years. Near or complete transmission elimination in dogs and humans was achieved after 10 years intervention using 50mg of pheromone under 70% household coverage in urban settings when newly acquired (immigrant) dogs had no pre-existing infections, but in rural settings required 90% coverage using 500mg. The willingness to pay (WTP) price per 10mg unit of pheromone is likely to be <$3 USD, making it a cost-beneficial intervention compared to current alternative strategies.

**Conclusions:** Integrated stochastic and spatial models capturing vector host-seeking behaviour, is a useful mathematical framework to evaluate spatially dependent intervention methods, fine-scale transmission dynamics, and to identify best practise.

## Introduction

Understanding arthropod vector behaviour is paramount to developing vector control tools against public and veterinary health diseases (Wilson 2020). *Leishmania* (Kinetoplastida: Trypanosomatidae) are intracellular protozoan parasites that primarily infect macrophages in the mammalian host causing human visceral leishmaniasis (VL), characterized by prolonged fever, wasting, splenomegaly, and hepatomegaly, with 90% case fatality in the absence of treatment (Martins-Melo 2014). Between 50,000 to 90,000 new human VL cases are reported per year (Alvar 2012; WHO 2017) occurring predominantly in East Africa, the Indian subcontinent, Central and South America, Mediterranean countries, and Central and Eastern Asia (Alvar 2012; Burza 2018).

*Leishmania* are transmitted between hosts by haematophagous female phlebotomine sand flies (Diptera: Psychodidae) (Torres-Guerrero et al., 2017). VL results from infection with *L. donovani* in an anthroponotic transmission cycle, or with *Le. infantum* in a zoonotic transmission cycle involving domestic dogs as the animal reservoir (Quinnell & Courtenay 2009). In dogs, *Le. infantum* infection often results in a multisystemic disease characterised by dermatitis, lymphadenomegaly, general muscular atrophy, and renal disease (Solano-Gallego 2011), being variably infectious to sand fly vectors (Courtenay 2014). This results in incidental “spill-over” transmission to humans. Humans and animals other than dogs, are considered non-reservoir (“dead-end”) hosts of *Le. infantum*, but nevertheless represent important blood sources supporting reproduction of the vector population (Alexander 2002).

Throughout the Americas, *Lutzomyia longipalpis* is the predominant sand fly vector of *Le. infantum*. The male sand fly releases a pheromone that mediates aggregation of male and female conspecifics at “leks” for the purposes of mating (Bray and Hamilton, 2007). Leks are usually located on or near to animal hosts which facilitates successful blood-feeding by female sand flies, resulting in *Le. infantum* transmission. The development of a synthetic copy of the male *Lu. longipalpis* sex-aggregation pheromone, produced from a low-cost plant derived intermediate (Bray et al., 2009), showed similar biological activity to the natural pheromone in attracting vectors to leks, and when placed in controlled-release dispensers, attracted vectors to co-located lethal pyrethroid insecticide (Bray et al., 2009; Bray et al., 2010; Bray et al., 2014). The synthetic pheromone, which appears to be dose-dependent (Bell et al., 2018), attracts females and other males from at least 30m from the source (Gonzales et al., 2020). Recent randomised control trials of this novel “lure-and-kill” approach in Brazil resulted in reductions of 52-53% in confirmed canine infection incidence and tissue *Le. infantum* parasite loads, and reductions of 49%-70% in vector abundance across treated households (Courtenay et al., 2019a; Gonçalves et al., in review). The impact was not dissimilar to a parallel trial arm where dogs were fitted with insecticide-impregnated collars, which have been documented to reduce human infection (Mazloumi et al., 2002) and clinical VL incidence when implemented under MoH operational conditions (Courtenay 2019b).

To date, the effectiveness of the “lure-and-kill” method to reduce human infection or clinical VL incidence has not been evaluated. To further understand the potential of this pheromone-based vector control tool, this study aimed to address outstanding questions, namely (i) what is the expected effect of the synthetic pheromone on sand fly vector attraction to human and animal hosts; (ii) what is the relationship between canine and human infection incidence; (iii) what are the most efficacious community-wide deployment strategies to reduce canine and human infection incidence under contrasting epi-demographical conditions; and (iv) what is the likely cost-effectiveness of the lure-and-kill tool. To answer these questions, we first quantified the spatial relationships between canine and human incidence in high (rural) and low (urban) transmission settings using data collected in Brazil.

The analytical framework combined a mathematical model for sand fly attraction to different host odour sources and synthetic pheromone; a stochastic spatially explicit model of transmission of *Le. infantum* between sand flies and dogs; and a mechanistic model of infection incidence in the human population. Detailed field and experimental data from high and low transmission settings in Brazil were available for model parameterisation. Simulations of the infection dynamics generated predictions of the relative impact of the lure-and-kill approach under variable deployment scenarios described by the quantity of synthetic pheromone deployed, the intervention coverage, the spatio-demographic population structure, and the infection status of immigrant dogs.

## Results

### Quantifying sand fly preference for hosts and pheromones

We conceptualized sand fly vector behaviour in response to sources of biochemical cues by constructing attraction profiles and parametrising them using aggregated data on *Lu. longipalpis* dispersal (Dye et al, 1991), host preference (Dilger, 2013), and capture success in synthetic pheromone baited light traps (Bell et al., 2018, Gonzales 2020) (Methods & Materials, SI Text). Figure 1 shows the predicted relative attraction of *Lu. longipalpis* to the most common blood-source hosts: chickens, dogs and humans, in the absence and presence of synthetic pheromone. The variability in the fraction of sand flies attracted to each source arises from the observed heterogeneity in host demography i.e. the numbers of humans, dogs and chickens, recorded in rural households (Dilger, 2013; Buckingham-Jeffery et al., 2018). Across households, in the absence of synthetic pheromone, a median 6.7% (95% C.I.s: 3.8%-36.6%) of sand flies are preferentially attracted to humans, 15.1% (95% C.I.s: 7.8%-84.0%) to dogs, and 81.2% (95% C.I.s: 54.4%-87.6%) to chickens. In the presence of 10mg synthetic pheromone controlled release dispensers, the expected median proportion of sand flies attracted to the three hosts decreased by approximately half: 3.1% (95% C.I.s: 2.2%-4.9%) (humans), 8.1% (95% C.I.s: 4.3%-22.2%) (dogs), and 42.1% (95% C.I.s: 14.8%-49.1%) (chickens) (Figure 1). Attraction to synthetic pheromone accounted for 53.7% (95% C.I.s: 39.7%-86.4%) of total *Lu. longipalpis* thus out-competing the host odours. This suggests that when co-located with insecticide, the lure-and-kill approach could dilute mean vector biting rates on humans and the canine reservoir by approximately half.

**Figure 1.**
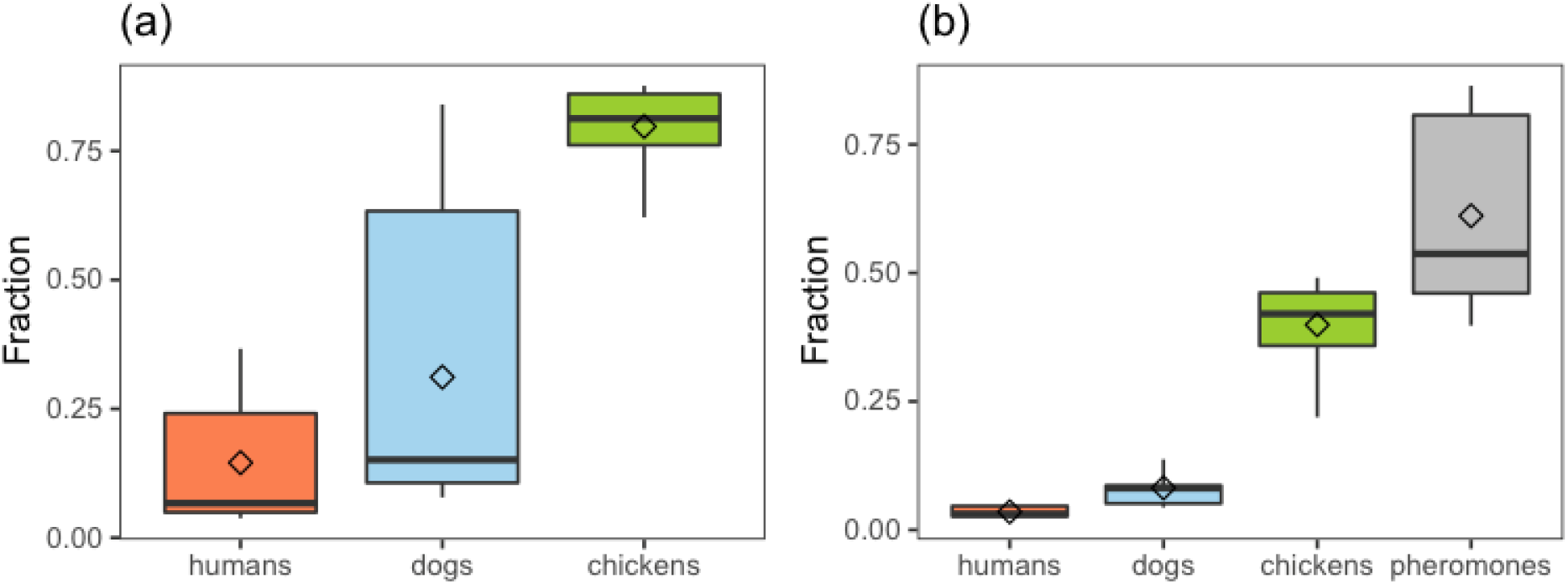
Estimated sand fly preference for hosts and pheromones. Attraction profiles were parametrised using the data on dispersal of *L*.*longipalpis* (Dye et al, 1991), host preference experiments (Dilger, 2013), and field experiments on sandfly capture in synthetic pheromone traps (González 2020; Bell et al., 2018) in Brazil. (a) Fraction of sand flies attracted to humans, dogs and chickens in each household were calculated based on demographic data in Caldeãro village, Pará state. (b) Those fractions were recalculated assuming that a trap with 10mg of synthetic pheromone was fitted for each household. Box plots show range (vertical line), interquartile range (box), median (horizontal line) and mean (diamonds).

### Spatial distributions of transmission rates in humans and dogs

Using the parametrized mathematical model and age-prevalence data for humans, the pre-intervention spatial distribution of the annual force of infection (FOI) in households were calculated (Figure 2; Supplementary materials; Supplementary Figure S11). For humans, the household FOI values ranged between 0.17 and 1.97 in the high transmission / rural setting, and between 0.013 and 0.037 in the low transmission / urban setting (Figure 2 (a)-(b)). Similarly, there was a high degree of variability in the annual FOI in household dogs, ranging from 0.05 to 17.1 in the rural setting, and from 0.02 to 0.53 in the urban setting (Figure 2 (c)- (d)). The FOI for rural dogs was over-dispersed (dispersion ratio d_r_= 2.364, p<0.001), while no overdispersion was found in the FOI for rural humans (d_r_=0.192, p=1), urban humans (d_r_=0.002, p=1) or urban dogs (d_r_=0.029, p=1). The post-control FOI for rural dogs was also over-dispersed, but at a smaller scale (d_r_=1.238, p=0.008). In the rural setting, the variation in the FOI values for dogs and humans were respectively 33.5× and 75× higher than in the urban settings. The variance in FOI values in dogs was 9.5× (rural) and 21× (urban) that in humans. Histograms of pre-control FOIs are shown in Figure 2 (e). These estimates are likely to reflect variations in the vector biting rates between hosts and households.

**Figure 2.**
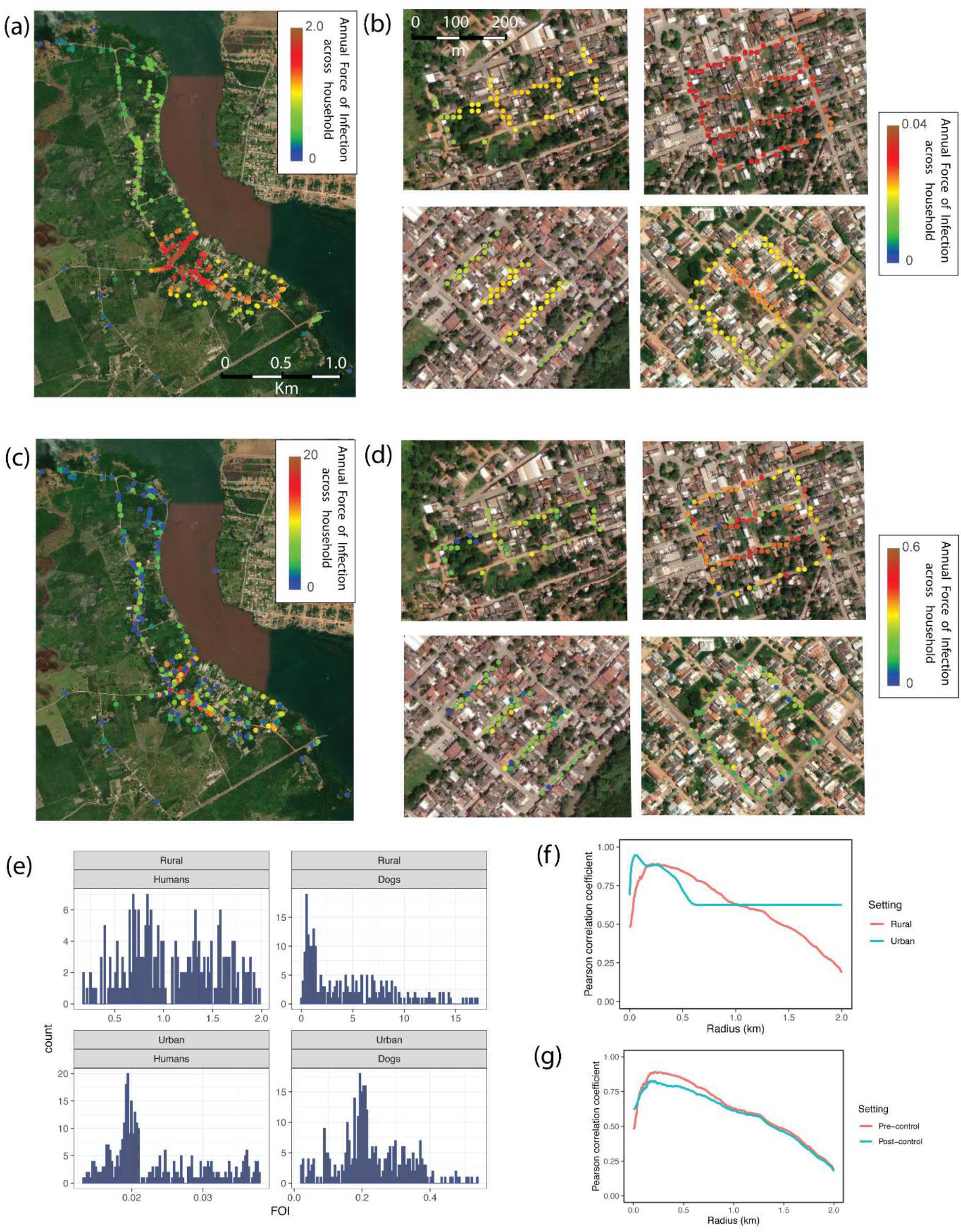
Distribution of force of infection (FOI). The spatial patterns of the annual pre-control distribution of force of infection for humans (a)-(b), and dogs (c)-(d). Here (a) and (c) is for the rural setting (Caldeãro village, Pará state), (b) and (d) for the urban settings (Governador Valadares, Minas Gerais state). For urban settings, each block is shown separately. Maps were drawn using the Wolfram Mathematica software [Mathematica12]. (e) Histograms of pre-control FOI. (f) Pearson correlation coefficient between pre-control FOI for humans and averaged FOI for dogs as a function of the radius. (g) Pearson correlation coefficient between pre-control ad post-control FOI for humans and averaged FOI for dogs as a function of the radius in rural setting and 50mg pheromone and 70% coverage.

The Moran’s I statistic for the FOI in humans were 0.63-0.82 (p<0.0001) in the four urban districts, and 0.56 (p<0.0001) in the rural setting, indicating significant spatial autocorrelation or clustering in both settings. By comparison, there was weak evidence for spatial clustering of FOI values in dogs in the urban settings (Moran’s I = 0.03-0.05, 0.036 ≤p≥ 0.064), but strong evidence of clustering in the rural setting (Moran’s I=0.12, p<0.0001). Pearson’s correlation coefficients were calculated between the FOI in humans and the average FOI in neighbouring canine households located within a specified radius of the human household; statistics were generated for 0 to 2km radii. The highest correlation corresponded to a radius of 220 meters distance from households in the rural setting (r_p_=0.89), and to 50 meters radius in the aggregated urban settings (r_p_=0.94) (Fig 2 (f)). The nearest-neighbour mean distance between rural houses was 40 meters, and between urban houses was 7 meters, indicating that the effective transmission radius from a house was between 5.5× (rural setting) and 7× (urban setting) the average distance between nearest-neighbours. Within households (radius equals 0 meters), the correlation coefficients were lower and not dissimilar between rural and urban settings (r_p_=0.48 and 0.69, respectively). Despite reductions in infection incidence attributed to the synthetic pheromone + insecticide intervention described below, the intervention did not significantly change this spatial relationship between human and dog transmission intensity. For example, in the rural setting, the shape of the relational curve pre-and post-intervention were similar (Fig 2g). The highest post-intervention correlation corresponded to a radius of 210 meters from households (r_p_=0.82) compared to 220 meters before control (Fig 2g).

### The impact of lure-and-kill implementation scenarios on infection

The short-term (1-2 years) and long-term (10 years) impacts of community-wide deployment of the lure-and-kill strategy on human and canine infection rates were assessed (Figure 3), varying the intervention coverage (percent of households per settlement receiving the intervention: 50%, 70% or 90%), and the amount of synthetic pheromone deployed at each household (10, 50, 100, or 500mg). All simulations were based on co-locating a constant quantity of insecticide.

**Figure 3.**
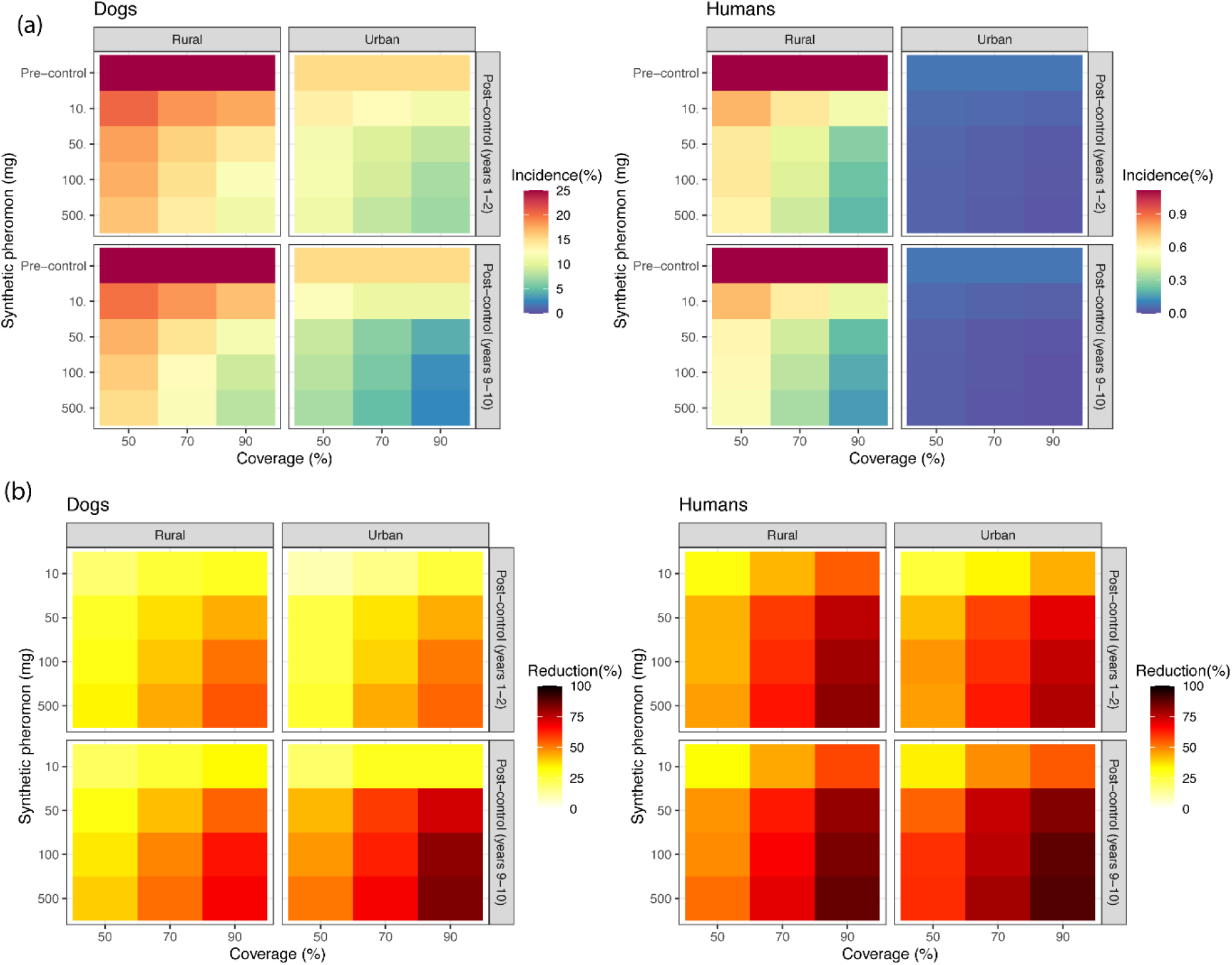
Reducing *L. infantum* transmission under the lure-and-kill approach: (a) annual incidence; (b) relative percentage reduction. Intervention coverage per settlement (percent of households with fitted pheromone + insecticide trap) was set to 50%, 70% and 90%, and the amount of synthetic pheromone placed in each lure dispenser was 10, 50, 100, and 500mg of synthetic pheromone. We assume that 13% of immigrant replacement dogs were exposed to *L*.*infantum* infection prior their introduction into the population. Mean incidence of new infections for humans is shown per 1000 persons per year.

The predicted reductions in canine and human incidence increased with the level of coverage and the amount of synthetic pheromone (Fig 3). Short-term reductions in human incidence were moderate to high (>50%), and not dissimilar in rural and urban settings, when deploying ≥50mg with ≥70% coverage (Figure 3b). Under the same intervention scenarios, the short term effect on canine infection incidence was more moderate (35%-65%), and differences between settings similarly not well pronounced (Figure 3b).

Following longer term interventions, human incidence was reduced substantially (>80%) in both rural and urban settings using 50mg lures with 90% coverage; or by >60% to 70% at a lower 70% coverage (Figure 3b). The relative impacts were greater than against canine incidence, which achieved >75% reductions only at 90% coverage using the larger amounts of pheromone in urban foci (Figure 3b). Under a regime of ≥70% coverage using 50mg pheromone, reductions in canine incidence of 66-78% compared to 47-59% were achieved in urban and rural foci respectively (Figure 3b). Thus overall, the lure-and-kill approach is generally predicted to be more effective under urban compared to rural conditions in the longer-term, and to be more efficient in reducing human than dog infection incidence.

### Relationship between the infection incidence in humans and dogs

Aggregating the results of the intervention scenarios, the annual infection incidence in humans showed a monotonic relationship with the annual incidence in dogs, in both rural and urban settings (Figure 4). The annual incidence in humans increased quadratically as the annual incidence in dogs increased. The implication is that controlling transmission to/from the reservoir has a disproportionate beneficial effect on human infection incidence. For example, in the rural setting, a 50% decrease in the annual canine incidence (from 24.4 pre-intervention to 12.2), is associated with a 71.7% drop (from 1.12 to 0.32) in human incidence (Fig 4a). Reducing the annual incidence in dogs by half in urban dogs resulted in a 59.9% reduction (from 0.078 to 0.031) in humans (Fig 4b). In this case, canine infection control has an 11.8% lower effectiveness against human incidence in low / urban compared to high / rural transmission settings, the difference being statistically significant (p<0.001).

**Figure 4.**
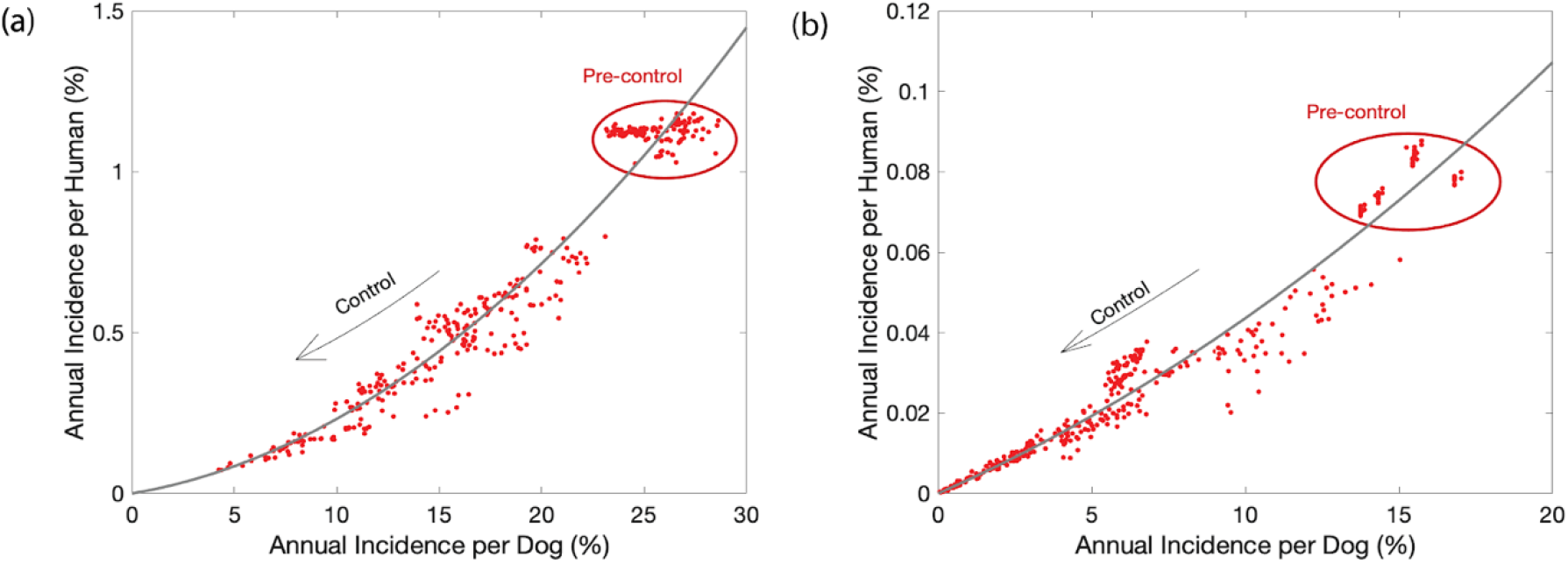
Relationship between annual incidence in dogs and humans: (a) rural setting; (b) urban setting. The line gives the fit of the exponential function.

### Effects of canine immigrant infection status on intervention success

The model simulations above assume that 13% of immigrant replacement dogs were exposed to *Le. infantum* infection prior their introduction into the population, as indicated by empirical observations. To assess the significance of imported canine infections, additional model simulations treated 100% of the immigrant dogs as susceptible (Supplementary Fig 10). The short-term (1-2 year) reductions in infection incidences in rural and urban settings were similar to the reductions achieved when a proportion of imported dogs were infectious (Supplementary figure 10). However, after continuous 10 years of 70% coverage and 50mg intervention, the model predicts reductions in urban infection incidence of up to 90% in both dogs and humans. In contrast, similar levels of reduction are achieved in the rural setting only under a regime of 90% coverage using 500mg of pheromone. Therefore, ensuring that newly acquired dogs are infection free can have a significant impact on the intervention efficacy.

### Cost-effectiveness of synthetic pheromone lures

The WTP unit price of the synthetic pheromone lure generally increased with the conversion ratio of human subclinical (asymptomatic) infection to VL clinical disease, leading to loss of DALYs (Figure 5). In rural settings for a conversion ratio of 1:6.5, the WTP price for 10mg of pheromone is between $2.03 (deployed at 50% coverage) and $2.40 (at 90% coverage). This compares to $0.72 and $0.84 respectively under a lower conversion ratio (1:18.5). In urban settings for a conversion ratio of 1:6.5, the WTP price for 10mg of pheromone is between $0.15 (at 50% coverage) and $0.16 (at 70% coverage). Considering an age-dependant structure of VL case numbers, substantially lower WTP unit prices ($0.32-$0.35) (rural) and ($0.021-$0.024) (urban) are indicated. In terms of WTP-defined thresholds of cost-effectiveness, the most cost-effective unit investment per DALY averted would be the deployment of a 10mg lure, under the expectation of 90% coverage in rural settings, and 70% coverage in urban settings.

**Figure 5.**
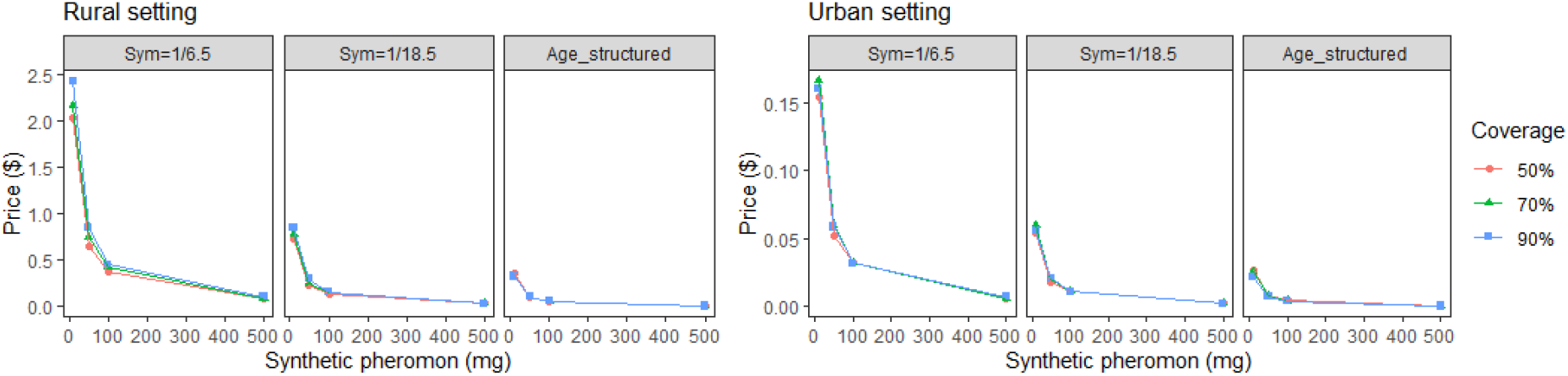
Estimated willingness to pay price for pheromone traps as a function of number of lures and coverage. The proportion of infections that are symptomatic was set to Sym=1/6.5 and Sym=1/18.5 (Rock et al., 2016); or the age-dependant proportion as defined in Davies and Mazloumi-Gavgani, 1999.

## Discussion

Previous mathematical models of *Leishmania* transmission and control have focused largely on non-spatial models (Rock et al., 2015). A novelty of this study is the integration of sub-models to formulate an individual-based, spatially explicit, VL transmission model. We simulated *Lu. longipalpis* attraction to different host odour sources and synthetic pheromone, transmission of *Le. infantum* between sand flies and dogs, which was coupled to a mechanistic model of infection incidence in the human population based on age-structured seroprevalence data. A key motivation was the need to provide a mathematical framework to optimally deploy interventions with spatial modes of action. In this study, we focussed on a novel lure-and-kill method which is the first time to our knowledge that a synthetic pheromone of an arthropod vector has been applied to control a public health disease (Bray et al., 2010; Courtenay 2019a; Goncalves in review).

The collective results suggest that for a given deployment scenario of the lure-and-kill method, it would be more effective against human than dog infection incidence, and with greater relative reductions achieved under low transmission / urban conditions than under high transmission / rural conditions, particularly in more sustained control programmes. This is because canine infection required either larger amounts of pheromone (50mg vs 500mg) or higher household coverages (50% vs 90%) to maintain similar reductions in higher compared to the transmission settings (Figure 3). However, considering the non-linear relationship between canine and human infection incidence (Figure 4), for a given reduction in canine infection incidence (50% for example), the impact on human infection incidence is expected to be less in urban than in rural settings (12% less in this example).

Unsurprisingly, increasing community-wide coverage to 90% and deploying 500mg of synthetic pheromone per household, presented the best case scenario towards reaching human VL elimination within a short time frame. Under a more realistic operational VL control programme target of 70% coverage, a 50mg pheromone lure + insecticide is expected to reduce rural human incidence by 58% (from 1.11 to 0.46) after 1-2 years, and by 70% (1.11 to 0.33) after 10 years. The same regime in urban areas reduced human incidence by 65% (0.076 to 0.026) after 1-2 years, and 73% (0.076 to 0.02) after 10 years. Similar short to long term patterns, but more moderate reductions, were predicted for canine infection incidence. The results suggest that the intervention effect is very strong at the start of the control programme, but gradually reduces in its proportional effect thereafter, signalling the need for sustained programmes to reach elimination.

This can be explained by the novel relationship revealed in canine and human transmission dynamics (Figure 3). The annual incidence in humans increased quadratically relative to the annual incidence in dogs in both transmission settings, but at different rates. Thus reductions in canine infection incidence do not map linearly onto reductions in human infection risk. As a consequence of this quadratic relationship, a higher gradient between reductions in canine and human incidence are observed when moving from pre-control to the early vs latter stages of control (Figure 4).

Empirical field trials of the lure-and-kill approach achieved a 52% reduction against parasite infection incidence in Brazilian dogs by deploying only 10mg synthetic pheromone + insecticide in households over 15 months (Courtenay 2019a). This suggests that the current model predictions may underestimate the true absolute effectiveness of the intervention. Nonetheless, the models are valuable in identifying the magnitude of the relative changes in transmission under varying demographic and intervention scenarios, and as shown for the first time here, the non-linear relationship between canine and human infection risk.

Model simulations also highlighted differences in the transmission dynamics between transmission / demographic settings, the FOI being a key measure of transmission potential (Keeling and Rohani, 2007). In the high transmission setting, the variation in the FOI values for dogs and humans were substantially higher than in urban settings, and higher in dogs than in humans in both settings, with significant over-dispersion especially in rural dogs. These estimates are likely to reflect differences in vector host choice and host accessibility, and in local vector biting rates. The protection attributed to the lure-and-kill intervention is illustrated by the observed reductions in household vector abundance in field trials (Courtenay 2019a; Goncalves, in review). In turn, this is expected to reduce the overdispersion in vector/host contact rates, with potential impacts on host parasite infection loads which influence their clinical development, infectivity and onward transmission (Courtenay 2017). The mean FOI was reduced 2-3 times post-control, although FOI distributions in both humans and dogs were of a similar shape post-intervention (Supplementary Figure 11). The additional over-dispersion observed in FOI values for rural dogs suggests that atypical pockets of enhanced transmission may occur and be more persistent in rural settings, thus requiring relatively higher levels of coverage to detect them.

The annual FOI in dogs (range: 0.04-17.1) in the rural setting is in line with the mean time to canine infection of 115 days (95% CI: 107-126 days) estimated previously in the same region (Quinnell 1997); the FOI (range: 0.019-0.53) estimates in the urban setting falls within the range of values calculated based on age-incidence-recovery modelled data from urban areas (Courtenay 2019a). In humans in the rural setting, approximately half of the household annual FOI values (range: 0.17-1.9) were FOI>1, indicating a higher relative infection pressure with a mean time from initial exposure to human infection being as little as 12 months.

Case clustering is a common feature of VL epidemiology (Courtenay 2017), and our spatial analysis detected clustering of households with high annual FOI values for humans (urban and rural) and dogs (rural only). Strongest spatial correlations between the FOI in humans and the average annual FOI amongst neighbouring dogs was at a 220 meter radius of rural houses, and 50 meter radius of urban houses. This reflects the considerably higher densities of houses and hosts in rural than urban settings which helps drive the differences in relative risk of infection between these settings.

The spatial clustering and dependence between human and dog infection rates is in general agreement with a number of observational studies in urban and rural settings across Brazil (Costa et al., 2018b; Campos et al., 2017; Araújo et al., 2013; Teixeira-Neto et al., 2014; Cavalho, et al., 2018; Dilger, 2013). Human infection risk associated with rising canine incidence has been reported by other modelling studies (Werneck et al., 2007), and canine surveys conducted in an urban setting (Araçatuba, São Paulo state), indicate spatial dependence of new and old canine infections on the density of infected dogs resident within 50m distance (Costa et al., 2018a). The parallel situation for anthroponotically transmitted *L. donovani* causing human VL in the Indian subcontinent, is illustrated by the increased risk of infection and VL in humans living with, or within similarly close proximity of, active VL case households (Chapman et al. 2018; Chapman et al., 2020). Within-household and/or near neighbour aggregation patterns are likely to arise in part due to the usual short range dispersal (<100m) of the respective sand fly vectors, measured by mark-release-recapture studies (Poché 2018; Dye 1991; Morrison 1993; Galvis-Ovallos 2018).

Simulating vector blood-seeking behaviour was particularly relevant in context of deploying a synthetic pheromone attractant. Determinants of host choice by blood-seeking vectors are multifactorial (Lehane, 2005). Feeding preferences by sand fly vectors have been modelled to depend on host defensiveness (Kelly, 2001), host irritability (Zahid and Kribs, 2020), host body surface area (Smith et al., 2016), and host species biomass (Buckingham-Jeffery et al., 2018). Other factors such as host accessibility are likely to be important (Agrela et al., 2002; Quinnell & Dye 1994; Courtenay 2007). The term attraction rate has been used in malaria models to quantify the blood-meal choice dependence on the propensity of hosts to emit mosquito attractants, and on host accessibility (Saul, 2003). In the current work, we introduce the term attraction profile to incorporate synthetic pheromone as a potential “choice” for sand flies. When constructing the attraction profiles, we assumed that attraction decreases exponentially with distance from a source, and a saturation effect is reached as the amount of synthetic pheromone in lures or numbers of hosts (kairomone quantity) increases (González et al., 2020; Bell et al., 2018; Kelly & Dye 1997). We model the localised influence of synthetic pheromone in treated households based on empirical data showing its attraction plume is at least 30 metres (González 2020), and that the attraction strength is non-linearly related to the pheromone quantity (dose-dependent) (Bell 2018). The proposed spatial positioning of the synthetic pheromone and insecticide within household compounds contrasts to that of experimental human odour lures to attract mosquitoes, which are longer range attractants, and therefore best placed further away from human residencies and other mosquito aggregation sites to reduce the risk of zoopotentiation (Okumu et al., 2010).

In Brazilian field studies, *Lu. longipalpis* occurs in highest abundance typically in chicken and other animal shelters rather than inside houses (Quinnell & Dye 1994; Alexander 2002). Keeping chickens is suggested to elicit zooprophylactic and zoopotentiation effects on infection incidence (Alexander 2002; Caldas et al., 2002; Belo 2013). Here, the modelling approach was to a combine these two possible effects such that the presence of chickens could elevate the numbers of vectors close to a household, but with a consequential increase of dilution of vector biting rates on humans and dogs. With or without pheromone treatment, humans were predicted to be the least attractive host, and chickens the most attractive host, to *Lu. longipalpis*. In the community trials of the lure-and-kill method, protection against canine infection, and decline in the local abundance of *Lu. longipalpis* vectors, were attributed to co-location of the synthetic pheromone and pyrethroid insecticide at domestic chicken roosting sites (Courtenay 2019a), with a greater effectiveness in households keeping chickens than those that did not (Goncalves In review). In agreement with those results, the model simulations here show a net zooprophylactic effect of the pheromone treatment across intervention scenarios. The synthetic pheromone does not inadvertently drive the vector onto the unprotected canine reservoir or humans (Courtenay 2019a), and keeping non-reservoir animal hosts, such as chickens, fowl and pigs, does not increase transmission risk, but rather can dilute biting rates on humans (Alexander 2002). However, keeping small livestock is not a prerequisite for the lure-and-kill method deployment, as the entomological field studies demonstrate (Gonçalves In review).

In line with sensitivity analyses in other modelling studies, the infection and infectiousness status of new dogs entering the population (immigration), in addition to vector components of vectorial capacity, significantly influenced VL model predictions (Buckingham-Jeffery et al., 2018). We assumed that 13% of newly introduced dogs were previously exposed and infectious, based on empirical observations in urban settings (Moreira et al. 2004). Estimate comparison against equivalent simulations where immigrant dogs were treated as susceptible, showed how the immigration of infectious dogs could lead to underestimates of the infection incidence (10-20% differences), and potentially undermine VL control programmes. A similar case has been suggested to cause mass anti-rabies vaccination programme failure in Chad (Laager et al., 2019).

The feasibility of a lure-and-kill strategy will also depend on economic evaluation being important for public policy decision making. Previous modelling of VL control options in Brazil suggest that IRS and insecticide-impregnated dog collars are the cheapest and/or the most cost-effective preventative strategies, being substantially lower than the costs associated with human VL case treatment (Shimozako et al, 2017). Well-designed benchmark cost-effective studies of VL control methods generally are lacking. Here we follow WHO’s guidelines basing cost–effectiveness thresholds on annual *per capita* gross domestic product (GDP) as a proxy of WTP per ability-adjusted life-year (DALY) averted (http://www.who.int/choice/en/ Accessed 8/2/20). The WTP unit price of synthetic pheromone increases with the subclinical to clinical VL conversion ratio. At face value, deployment of a 10mg pheromone lure assuming 90% coverage in rural and urban areas, achieves the greatest cost-effectiveness in terms of WTP-defined thresholds. Under the more realistic target of 70% coverage, the WTP values are not that dissimilar. Under the observed age-dependant VL case structures (Hayhay 2011B; Davies and Mazloumi-Gavgani, 1999; Martins Melo et al, 2018), all scenarios tested here predict a unit WTP of less than US$1. These costs are low relative to common vector control methods. For example, the mean WTP for insecticide-treated nets to combat malaria estimated by meta-analysis was US$ 2.79 (95% CL: 1.07, 4.51) (Trapero-Bertran et al., 2013). For simplicity, we calculated WTP based on estimated synthetic pheromone lure unit manufacturing costs; implementation costs were not included but are reasonably assumed to be similar to delivery of IRS as a reference. In this respect, the costs of the lure-and-kill deployment may be offset against a >90% saving in insecticide requirements for IRS (Goncalves In review). A more complete economic assessment is now needed in context of defined health-benefits and expenditure targets (Marseille et al 2015).

More than 95% of the 11,049 VL incident cases reported in the Americas between 2015-2017 occurred in Brazil (PAHO 2019). The age-standardized human VL incidence, and years of life lost (YLL), have increased in Brazil since 1990 by 53% and 108% respectively, and particularly so in young (0-4 year old) children (Bezerra et al., 2018; Martins Melo et al, 2018). The Brazilian national VL control guidelines promote human case detection and treatment, and IRS on detection of a new VL case (MOH 2014; Camargo-Neves 2006). There is no human vaccine (Iborra et al., 2018). To prevent transmission from dogs, the program recommends euthanasia of seropositive dogs, however this is unpopular leading to low compliance (Zuben & Donalísio 2016). Furthermore, dogs lost to the population through all-cause mortality (increased by culling) are replaced by dog-owners such that the canine population is stationary and approximately stable (Courtenay 1998). Canine vaccination, topical insecticides and therapeutic treatments are also promoted, but variably adopted or effective in reducing transmission (MOH 2014; Dantas-Torres 2020; Miro 2017; Nogueira 2019). That transmission persists (Shimozako 2017; PAHO 2019; da Rocha et al., 2018), and its range is expanding from rural into urban settings, and across national and international borders (PAHO 2019; Harhay 2011A; Seva 2017), is a strong motivation to develop sustainable preventative vector control methods.

In summary, this study highlights how canine infection drives human infection incidence, and the differences in transmission dynamics and spatial relationships in contrasting transmission and demographic settings. To achieve similar impacts on human incidence across these settings may require greater programmatic investment in high incidence / rural versus in urban settings, as control of canine incidence may have a proportionally lower impact on human infection. Amongst realistic intervention scenarios, such as 70% coverage using 50mg pheromone, the lure-and-kill method is predicted to be both efficacious and highly cost-effective in rural and urban settings. The integration of mathematical models to capture key components of transmission including vector behavioural responses, is a useful framework to evaluate intervention methods with spatial modes of action. As voiced by this and previous studies (Wilson 2020; Okuma 2010), successful manipulation of vectors to control transmission requires a good understanding of vector behaviour, including their responses to both host- and vector-mediated aggregation cues.

## Materials and Method

### Mathematical model

A stochastic spatial model was developed to incorporate four processes: (i) distribution of sand flies across a heterogeneous landscape created by host kairomone and vector pheromone attraction profiles; (ii) transmission of *Le. infantum* between dogs and sand flies; (iii) transmission of *Le. infantum* from sand flies to humans; and (iv) the effects of a synthetic pheromone + insecticide intervention against transmission. The model is described below. More details are provided in the Supplementary Information.

### Attraction profiles for hosts and pheromones

The local distribution of sand flies was constructed in terms of attraction profiles. The attraction profile for a household accounts for the presence of hosts (humans, dogs and chickens) and synthetic pheromones lures. We have parametrised attraction profiles using the data on dispersal of *Lu. longipalpis* (Dye et al, 1991), host-biting preference (Dilger, 2013), and sand fly capture success in synthetic pheromone baited field traps (Bell et al., 2018; Gonzales 2020). We assumed that the strength of the pheromone attraction decreases with the distance from the lure with an exponential decay, and that there is a saturation effect with increasing quantity of pheromone deployed (Bell 2018), or number of hosts. The probability that a sand fly will be attracted to a source is found by the integration of the convolution between the attraction profile and the dispersal kernel. We used the MCMC algorithm to estimate the parameters from the 309 available data sets (Roberts and Rosenthal, 2009) (Supplementary Information; Supplementary Figures S2-S4).

### Transmission of visceral leishmaniasis between dog and sand flies

The dog population comprised five infection states: susceptible, exposed, never infectious, of low infectiousness, and of high infectiousness, with defined periods of latency (Courtenay 2014). Each dog at the time of introduction into the population was randomly assigned to one of the three infectious states in fractions specified by field observations: probability of high infectiousness equal to 0.37 and probability of low infectiousness equal to 0.08 (Courtenay et al., 2002; Courtenay et al., 2014). Dogs were removed due to a death only, assuming exposed and infectious dogs have higher mortality rates compared to susceptible dogs. Deceased dogs were replaced by newly introduced dogs, which have a probability of being already infected. The VL transmission model is based on a localised vectorial capacity of the sand fly population transmitting infection between dogs [Dye, 1996; Gomez et al., 2018]. We modified the force of infection (FOI) to include a spatial dependence on a dispersal kernel function between infectious and susceptible dogs (Supplementary Information).

### Model parametrisation

Models were parameterised for contrasting epidemiological and demographic conditions in endemic Brazil: the rural setting was characterised by low household and host population densities/m^2^, and high transmission rates based on data collected in Marajó, Pará state, in north Brazil (Dilger, 2013; Courtenay et al., 2002; 2014; Dye et al, 1991; 1996). The urban setting was characterised by higher household and host densities/m^2^, and low transmission rates, using data collected in municipalities in Aracatuba, Sao Paulo state (Courtenay 2019a) and Governador Valadares, Minas Gerais state (Bell 2018; Gonzales 2020; Gonçalves, In review) (Figure 2; Methods & Materials, SI Text).

As a measure of relative sand fly vector abundance through calendar time, we used the number of sand flies captured per trap night in rural Marajó (Dilger, 2013), incorporating a sinusoidal function fitted to these temporal data (Supplementary Information; Supplementary Figure S5).

The posterior distribution of sand fly absolute scaling was based on canine infection prevalence in Marajó (Dilger, 2013), estimated by approximate Bayesian computation using a sequential Monte Carlo method (Minter and Retkute, 2019). The distance metrics represent absolute differences between observed infection prevalence in dogs and simulated infection prevalence in dogs 10 years after infected case seeding. The observed infection prevalence in dogs was initially set at 49.3% (Courtenay 2019a). We compared the number of *Lu. longipalpis* captured by CDC light traps in Marajó to the number captured in Araçatuba city (Dilger, 2013). The ratio between the two settings being 0.028 (Supplementary Figure S6). Therefore, for simulations in urban setting we further rescaled the abundance of sand flies by 0.028.

The posterior distribution of the scaling parameter and simulated infection prevalence in dogs represented 100 replicate values sampled from the poster distribution, are shown in Supplementary Figure S7.

Additional model parameters were obtained from published literature (Courtenay et al., 2002, 2014; Dye et al, 1991; Dye, 1996; Moreira et al., 2004; Nunes et al., 2008; Reithinger et al., 2004; Ribas et al., 2013; Shimozako et al. 2017) listed in Supplementary Table 2. *Le. infantum* transmission was simulated in dogs and humans in rural and urban settings. The pre-control equilibrium prevalence in dogs was 40%-60% in the rural setting, and 20%-40% in the urban setting (Supplementary Figure S9).

### Estimating seroprevalence in the human population

The probability of infection with age was calculated based on previously described models (Courtenay et al., 1994; Williams and Dye, 1994):

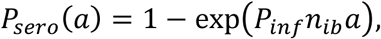

where *a* is age, *P*_*inf*_ is the probability of becoming infected after being bitten by an infected sand fly; *n*_*ib*_ is the number of infectious bites per human host. We have estimated *P*_*inf*_ from the age-exposure data in humans from Marajó villages, and using simulated values of number of infectious bites (Supplementary Information; Supplementary Figure S8).

### Computing spatial autocorrelation and over-dispersal of FOI

The Moran’s I statistic was calculated in R (package *spdep*) (Bivand et al., 2018). Neighbourhood contiguity was calculated from Euclidean distance between coordinates of households and taking bounds between 0 and 250 meters. The Moran’s I statistic evaluates whether the pattern is clustered, dispersed, or random.

We used R packages *lme4* and *performance* first to fit generalized linear models in the form *FOI∼humans + dogs + chicken* (Bates et al., 2015), and then to run overdispersal test (Lüdecke et al., 2020). Dispersion ratios larger than one indicate overdispersion.

### Intervention scenarios

Model simulations were initiated by introduction of a single infectious dog and allowed to run for 10 years until canine infection prevalence reached endemic equilibrium. The intervention, pheromone lures and insecticide, were then introduced, considering the following implementation regimes: coverage (% of households treated) 50%, 70% and 90%; and synthetic pheromone quantity: 10mg, 50mg, 100mg and 500mg. Each house received pheromone slow-release dispensers which were assumed to be replaced sufficiently frequently (Gonzales et al., 2020). Simulations were run for an additional 10 years after the introduction of the interventions. (Supplementary Information; Supplementary Figure S10). We also investigated two scenarios for replacement dogs: (i) import of infection where 13% were considered exposed prior to immigration (Moreira et al. 2004, Nunes et al. 2008); and (ii) no import of infection.

Computer code was written in R (https://CRAN.R-project.org) and is available from https://github.com/rretkute/VLsnthPherModel.

### Estimating willingness to pay (WTP)

One financial indicator of the cost-effectiveness of a control strategy is the Willingness to Pay (WTP), which is the monetary value of a unit disability-adjusted life-year (DALY) averted; the monetary threshold to consider an intervention cost–effective is related to annual *per capita* gross domestic product (GDP) (Marseille 2015; Brouwer and Bateman 2005). Here we follow WHO’s guidelines that one year of life is worth 50% of *per capita* GDP (http://www.who.int/choice/en/ Accessed 8/2/20). WTP calculations here accounted for: (i) simulated number of infections averted by implementation of control options; (ii) the proportion of infections that were symptomatic VL cases; (iii) case fatality rate (CFR); (iv) years of life lost due to premature death (YLL); and (v) the *per capita* GDP. Average annual age-adjusted CRF was estimated as 8.1% (Martins-Melo et al., 2014). The standard life expectancy at birth is 86.6 years (Bezerra et al., 2018), and we assumed GDP=$8,717 for Brazil (Word Bank, 2019). We have not included in the analysis years lost due to disability, as it makes a negligible contribution to total DALYs for VL (Heydarpour et al., 2016).

The ratio of subclinical (asymptomatic) human infections to clinical VL disease leading to loss of DALYs varies globally (Rock et al., 2016). Here, WTP was calculated for two ratios: 1/6.5 and 1/18.5. We also calculated WTP for an age-dependent distribution in VL incidence where the highest proportion (0.15) of infections causing VL are in children 1-3 years old, and a negligible proportion are in the 8+ year old age groups (Davies and Mazloumi-Gavgani, 1999). WTP was calculated for 50%, 70% and 90% intervention coverage for urban (low transmission, high host densities/m^2^) and rural (high transmission, low host densities/m^2^) settings.

## Data Availability

https://github.com/rretkute/VLsnthPherModel

## Acknowledgments

The authors acknowledge the multiple collaborations that have led to the generation of the referenced data used in this study. We also thank members of the Zeeman Institute, University of Warwick, for helpful discussions during the preparation of this manuscript.

## Data availability

The data supporting the conclusions of this article are included within the article. Data are available from the corresponding authors on reasonable request to be used solely within the context of this study, following the ethical agreements, and with permission from the relevant authorities and co-authors.

## References

Agrela I., Sanchez E., Gomez B., Feliciangeli M.D. (2002) Feeding behavior of Lutzomyia pseudolongipalpis (Diptera: Psychodidae), a putative vector of visceral leishmaniasis in Venezuela. J Med Entomol. 39(3): 440–5.

Alexander B., Lopes de Carvalho R., McCallum H., Pereira M.H. (2002) Role of the domestic chicken (Gallus gallus) in the epidemiology of urban visceral leishmaniasis in Brazil Emerg. Infect. Dis., 8: 1480–1485.

Alvar, J., V’elez, I. D., Bern, C., Herrero, M., Desjeux, P., Cano, J., Jannin, J., and den Boer and, M. (2012). Leishmaniasis worldwide and global estimates of its incidence. PLoS ONE, 7(5):e35671.

Araújo V.E.M., Pinheiro L.C., Mattos Almeida M.C., Menezes F.C., Morais M.H.F., Reis I.A., Assunção R.M., Carneiro M. (2013) Relative Risk of Visceral Leishmaniasis in Brazil: A Spatial Analysis in Urban Area. PLoS Negl Trop Dis. 7(11) e2540.

Assis T.S.M., Rabello A., Cota G., Werneck G.L., Azeredo-da-Silva ALF. (2019) Cost-effectiveness analysis of diagnostic-therapeutic strategies for visceral leishmaniasis in Brazil.. Rev Soc Bras Med Trop, 52:e20180272.

Baneth, G., Koutinas, A. F., Solano-Gallego, L., Bourdeau, P., and Ferrer, L. (2008). Canine leishmaniosis – new concepts and insights on an expanding zoonosis: part one. Trends in Parasitology, 24(7):324–330.

Bates D, Mächler M, Bolker B, Walker S (2015). “Fitting Linear Mixed-Effects Models Using lme4.” Journal of Statistical Software, 67(1), 1–48. doi:10.18637/jss.v067.i01.

Bell, M. J., Sedda, L., Gonzalez, M. A., de Souza, C. F., Dilger, E., Brazil, R. P., Courtenay, O., and Hamilton, J. G. C. (2018). Attraction of lutzomyia longipalpis to synthetic sex-aggregation pheromone: Effect of release rate and proximity of adjacent pheromone sources. PLOS Neglected Tropical Diseases, 12(12):e0007007.

Bermudi P.M.M., Guirado M.M., Rodas L.A.C., Dibo M.R., Chiaravalloti-Neto F. (2018) Rev Soc Bras Med Trop. 2018 Jul-Aug; 51(4):452–460. doi: 10.1590/0037-8682-0505-2017. Spatio-temporal analysis of the occurrence of human visceral leishmaniasis in Araçatuba, State of São Paulo, Brazil.

Bezerra, J. M. T., de Arau’jo, V. E. M., Barbosa, D. S., Martins-Melo, F. R., Werneck, G. L., and Carneiro, M. (2018). Burden of leishmaniasis in brazil and federated units, 1990-2016: Findings from global burden of disease study 2016. PLOS Neglected Tropical Diseases, 12(9):e0006697.

Bivand R, Wong DWS (2018). Comparing implementations of global and local indicators of spatial association. TEST, 27(3), 716–748.

Braga G.B., Remoaldo P.C., de Carvalho Fiúza A.L. (2016) A methodology for definition of rural spaces: an implementation in Brazil. Cienc. Rural 46(2) 0103-8478cr20150464.

Bray, D. P. and Hamilton, J. G. C. (2007). Host odor synergizes attraction of virgin female (diptera: Psychodidae). Journal of Medical Entomology, 44(5):779–787.

Burza, S., Croft, S. L., and Boelaert, M. (2018). Leishma-niasis. The Lancet, 392(10151):951–970.

Bray, D. P., Bandi, K. K., Brazil, R. P., Oliveira, A. G., and Hamilton, J. G. C. (2009). Synthetic sex pheromone attracts the leishmaniasis vector (diptera: Psychodidae) to traps in the field. Journal of Medical Entomology, 46(3):428–434.

Bray, D.P., Alves, G.B., Dorval, M.E., Brazil, R.P. and Hamilton, J.G.C. (2010) Synthetic sex pheromone attracts the leishmaniasis vector Lutzomyia longipalpis to experimental chicken sheds treated with insecticide. Parasites & Vectors 3, 16.

Bray, D. P., Carter, V., Alves, G. B., Brazil, R. P., Bandi, K. K., and Hamilton, J. G.C. (2014). Synthetic sex pheromone in a longlasting lure attracts the visceral leishmaniasis vector, lutzomyia longipalpis, for up to 12 weeks in brazil. PLoS Neglected Tropical Diseases, 8(3):e2723.

Brouwer, R., and Bateman, I.J. 2005. Benefits Transfer of Willingness to Pay Estimates and Functions for Health-Risk Reductions: A Cross-Country Study. Journal of Health Economics 24 (3): 591–611.

Buckingham-Jeffery E., Hill E.M., Datta S., Dilger E., Courtenay O. (2019) Spatio-temporal modelling of Leishmania infantum infection among domestic dogs: a simulation study and sensitivity analysis applied to rural Brazil. Parasit Vectors 12(1): 215.

Caldas A.J.M., Costa J.M.L., Silva A.A.M., Vinhas V., Barral A. (2002) Risk factors associated with asymptomatic infection by Leishmania chagasi in north-east Brazil Trans. R. Soc. Trop. Med. Hyg., 96 21–28.

Camargo-Neves V.L.F., Glasser C.M., Cruz L.L., de Almeida R.G. (2006) Manual de Vigilancia e Control da Leishmaniose Visceral Americana de Estado de Sao Paulo. Sao Paulo.

Campos R., Santos M., Tunon G., Cunha L., Magalhães L., Moraes J., Ramalho D., Lima S., Pacheco J.A., Lipscomb M., Ribeiro de Jesus A., Pacheco de Almeida R. (2017) Epidemiological aspects and spatial distribution of human and canine visceral leishmaniasis in an endemic area in northeastern Brazil. Geospat Health 12(1) 503.

Carvalho A.G., Luz J.G.G., Rodrigues L.D., Dias J.V.L., Fontes C.J.F. (2018) High seroprevalence and peripheral spatial distribution of visceral leishmaniasis among domestic dogs in an emerging urban focus in Central Brazil: a cross-sectional study. Pathog Glob Health 112(1) 29–36.

Chapman L.A.C., Jewell C.P., Spencer S.E.F.1, Pellis L., Datta S., Chowdhury R., Bern C., Medley G.F., Hollingsworth T.D. (2018) The role of case proximity in transmission of visceral leishmaniasis in a highly endemic village in Bangladesh. PLoS Negl Trop Dis. 12(10) e0006453.

Chapman L.A.C, Spencer S.E.F, Pollington T.M, Jewell C.P., Mondal D., Alvar J., Hollingsworth T.D, Cameron M.M, Bern C., Medley G.M. (2020) Inferring transmission trees to guide targeting of interventions against visceral leishmaniasis and post-kala-azar dermal leishmaniasis. Proc Natl Acad Sci U S A. 202002731.

Costa C.H. (2008) Characterization and speculations on the urbanization of visceral leishmaniasis in Brazil. Cad Saude Publica 24(12) 2959–63.

Costa, D.N.C.C., Code,co, C.T., Silva, M.A., and Werneck, G.L. (2013). Culling dogs in scenarios of imperfect control: Realistic impact on the prevalence of canine visceral leishmaniasis. PLoS Neglected Tropical Diseases, 7(8):e2355.

Costa D.N.C.C., Blangiardo M., Rodas L.A.C., Nunes C.M., Hiramoto RM5, Tolezano JE5, Bonfietti LX6, Bermudi PMM1, Cipriano RS7, Cardoso GCD7, Codeço C.T., Chiaravalloti-Neto F. (2018) Canine visceral leishmaniasis in Araçatuba, state of São Paulo, Brazil, and its relationship with characteristics of dogs and their owners: a cross-sectional and spatial analysis using a geostatistical approach. BMC Vet Res. 14(1) 229.

Costa D.N.C.C., Bermudi P.M.M., Rodas L.A.C., Nunes C.M., Hiramoto R.M., Tolezano J.E., Cipriano R.S., Cardoso G.C.D., Codeço C.T., Chiaravalloti Neto F. (2018) Human visceral leishmaniasis and relationship with vector and canine control measures. Rev Saude Publica. 52 92.

Courtenay O., Macdonald D.W., Lainson R., Shaw J.J., Dye C. (1994) Epidemiology of canine leishmaniasis: a comparative serological study of dogs and foxes in Amazon Brazil. Parasitology 109 273–9.

Courtenay, O., Quinnell, R. J., Garcez, L. M., Shaw, J. J., and Dye, C. (2002). Infectiousness in a cohort of brazilian dogs: Why culling fails to control visceral leishmaniasis in areas of high transmission. The Journal of Infectious Diseases, 186(9): 1314–1320.

Courtenay O., Carson C., Calvo-Bado L., Garcez L.M., Quinnell R.J. (2014) Heterogeneities in leishmania infantum infection: Using skin parasite burdens to identify highly infectious dogs. PLoS Neglected Tropical Diseases 8(1): e2583.

Courtenay O., Peters N.C., Rogers M.E., Bern C. (2017) Combining epidemiology with basic biology of sand flies, parasites, and hosts to inform leishmaniasis transmission dynamics and control. PloS Pathogens 13(10): e1006571.

Courtenay O., Dilger E., Calvo-Bado L.A., Kravar-Garde L., Carter V., Bell M.J., Alves B.J., Goncalves R., Makhdoomi M.M., González M.A., Nunes C.M., Bray D.P., Brazil R.P., Hamilton J.G.C. (2019) Sand fly synthetic sex-aggregation pheromone colocated with insecticide reduces the incidence of infection in the canine reservoir of visceral leishmaniasis: A stratified cluster randomised trial. PLoS Negl Trop Dis 13(10) e0007767.

Courtenay O., Bazmani A., Parvizi P., Ready P.D., Cameron M.M. (2019) Insecticide-impregnated dog collars reduce infantile clinical visceral leishmaniasis under operational conditions in NW Iran: A community-wide cluster randomised trial. PLoS Negl Trop Dis. 13(3): e0007193.

Cucunubá Z.M., Nouvellet P., Conteh L., Vera M.J., Angulo V.M., Dib J.C., Parra-Henao G.B., Basáñez M.G. (2017) Modelling historical changes in the force-of-infection of Chagas disease to inform control and elimination programmes: application in Colombia. BMJ Glob Health 2 e000345.

da Rocha, I. C. M., dos Santos, L. H. M., Coura-Vital, W., da Cunha, G. M. R., do Carmo Magalh aes, F., da Silva, T. A. M., Morais, M. H. F., Oliveira, E., Reis, I. A., and Carneiro, M.(2018). Effectiveness of the brazilian visceral leishmaniasis surveillance and control programme in reducing the prevalence and incidence of leishmania infantum infection. Parasites & Vectors, 11(1).

Dantas-Torres, F. and Brand∼ao-Filho, S. P. (2006). Visceral leishmaniasis in brazil: revisiting paradigms of epidemiology and control. Revista do Instituto de Medicina Tropical de S ∼ao Paulo, 48(3):151–156.

Vaccination against canine leishmaniasis in Brazil. Int J Parasitol. In Press

Davies C.R., Mazloumi-Gavgani A.S. (1999) Age, acquired immunity and the risk of visceral leishmaniasis: a prospective study in Iran. Parasitology, 119 (Pt 3):247–57.

de Carvalho I.P.S.F., Peixoto H.M., Romero G.A., de Oliveira M.R.F. (2019) Treatment for human visceral leishmaniasis: a cost-effectiveness analysis for Brazil. Trop Med Int Health, 24(9): 1064–1077.

Dye C., Davies C.R., Lainson R. (1991) Communication among phlebotomine sandflies: a field study of domesticated lutzomyia longipalpis populations in amazonian Brazil. Animal Behaviour, 42(2): 183–192.

Dye C. (1996). The logic of visceral leishmaniasis control. The American Journal of Tropical Medicine and Hygiene, 55(2): 125–130.

Dilger, E. (2013). The effects of host-vector relationships and density dependence on the epidemiology of visceral leishmaniasis. PhD thesis, University of Warwick.

Fernández M.S., Salomón O.D., Cavia R., Perez A.A., Guccione J.D. (2010) Lutzomyia longipalpis spatial distribution and association with environmental variables in an urban focus of visceral leishmaniasis, Misiones, Argentina. Acta Trop. 114 81–87.

Galvis-Ovallos F., Casanova C., Bergamaschi D.P., Galati E.A.B. (2018) A field study of the survival and dispersal pattern of Lutzomyia longipalpis in an endemic area of visceral leishmaniasis in Brazil. Plos Neglected Tropical Diseases 12(4): e0006333.

Gomez S.A., Chapman L.A.C., Dilger E., Courtenay O., Picado A. (2018) Estimating the efficacy of community-wide use of systemic insecticides in dogs to control zoonotic visceral leishmaniasis: A modelling study in a brazilian scenario. PLOS Neglected Tropical Diseases, 12(9):e0006797, sep 2018.

González MA, Bell M, Souza CF, Freitas RM, Brazil RP, Courtenay O, Hamilton JGC. Synthetic sex-aggregation pheromone of Lutzomyia longipalpis, the South American sand fly vector of Leishmania infantum, attracts males and females over long-distance. PLoSNTDs In Press.

Hamilton J.G. (2008) Sandfly pheromones. Their biology and potential for use in control programs. Parasite 15(3) 252–6.

Harhay M.O., Olliaro P.L., Costa D.L., Costa C.H. (2011) Urban parasitology: visceral leishmaniasis in Brazil. Trends Parasitol 27(9) 403–9.

Harris M., Caldwell J.M., Mordecai E.A. (2019) Climate drives spatial variation in Zika epidemics in Latin America. Proc Biol Sci. 286(1909) 20191578.

Heydarpour F., Sari A.A., Mohebali M., Shirzadi M., Bokaie S. (2016) Incidence and Disability-Adjusted Life Years (Dalys) Attributable to Leishmaniasis In Iran, 2013. Ethiop J Health Sci, 26(4): 381–8.

Hollingsworth T.D. (2018) Counting Down the 2020 Goals for 9 Neglected Tropical Diseases: What Have We Learned From Quantitative Analysis and Transmission Modeling? Clin Infect Dis. 66 (suppl_4) S237–S244.

Iborra, S., Solana, J. C., Requena, J. M., and Soto, M. (2018). Vaccine candidates against leishmania under current research. Expert Review of Vaccines, 17(4):323–334.

Keeling M.J., Rohani P. (2007) Modeling Infectious Diseases in Humans and Animals. Princeton University Press.

Kelly D.W. (2001) Why are some people bitten more than others? Trends Parasitol. 17(12) 578–81.

Laager M., Léchenne M., Naissengar K., Mindekem R., Oussiguere A., Zinsstag J., Chitnis N. (2019) A metapopulation model of dog rabies transmission in N’Djamena, Chad. J Theor Biol. 462 408–417.

Lainson R., Rangel E.F. (2005) Lutzomyia longipalpis and the ecoepidemiology of American visceral leishmaniasis, with particular reference to Brazil: a review Mem. Inst.Oswaldo Cruz, 100 (2005), pp. 811–827

Lehane M.J. (2005) The Biology of blood-sucking insects. Cambridge University Press.

Lima I.D., Lima A.L.M., Mendes-Aguiar C.D., Coutinho J.F.V., Mary E.W., Pearson R.D., (2018) Changing demographics of visceral leishmaniasis in northeast Brazil: Lessons for the future. Plos Neglected Tropical Diseases 12(3) e0006164.

Lima, A’. L.M., deLima, I.D., Coutinho, J.F.V., deSousa, U. P. S. T., Rodrigues, M. A. G.,Wilson, M. E., Pearson, R. D., Queiroz, J. W., and Jer^onimo, S. M. B. (2017). Changing epidemiology of visceral leishmaniasis in northeastern brazil: a 25-year follow-up of an urban outbreak. Transactions of The Royal Society of Tropical Medicine and Hygiene, 111(10):440–447.

Lüdecke D, Makowski D, Waggoner P, Patil I (2020). “performance: Assessment of Regression Models Performance.” CRAN. doi: 10.5281/zenodo.3952174

Marseille E., Larson B., Kazi D.S., Kahn J.G., Rosen S. (2015) Thresholds for the cost–effectiveness of interventions: alternative approaches. Bull World Health Organ 2015;93:118–124. doi: http://dx.doi.org/10.2471/BLT.14.138206

Martins-Melo F.R., Lima Mda S., Ramos A.N.Jr., Alencar C.H., Heukelbach J. (2014) Mortality and case fatality due to visceral leishmaniasis in Brazil: a nationwide analysis of epidemiology, trends and spatial patterns. PLoS One, 9(4): e93770.

Martins-Melo F.R., Carneiro M., Ramos N.V, Heukelbach J., Ribeiro A.L.P., Werneck G.L. (2018) The burden of Neglected Tropical Diseases in Brazil, 1990-2016: A subnational analysis from the Global Burden of Disease Study 2016. PLoS Neglected Tropical Diseases 12(6) e0006559.

Wolfram Research, Inc., Mathematica, Version 12.0, Champaign, IL (2019).

Menon, S. S., Rossi, R., Nshimyumukiza, L., and Zinszer, K. (2016). Decentralized control of human visceral leishmaniasis in endemic urban areas of brazil: a literature review. Tropical Medicine and Health, 44(1).

Minter A., Retkute R. (2019) Approximate Bayesian computation for infectious disease modelling. Epidemics, 29 100368.

Ministério da Saúde B. Manual de vigilância e controle da leishmaniose visceral. In: Epidemiológica SdVeSDdV, editor. 1 ed. Brasília: Ministério da Saúde.; 2014. p. 120.

Moreira E.D., de Souza V.M.M., Sreenivasan M., Nascimento E.G., de Carvalho L.P. (2004) Assessment of an optimized dog-culling program in the dynamics of canine leishmania transmission. Veterinary Parasitology 122(4) 245–252.

Morrison A.C., Ferro C., Morales A., Tesh R.B., Wilson M.L. (1993) Dispersal of The Sand Fly Lutzomyia-Longipalpis (Diptera, Psychodidae) at an Endemic Focus of Visceral Leishmaniasis in Colombia. Journal of Medical Entomology 30(2): 427–35.

Nunes C.M., de Lima M.V.F., de Paula H.B., Perri S.H.V., de Andrade A.M., Dias F.E.F., Burattini M.N. (2008) Dog culling and replacement in an area endemic for visceral leishmaniasis in Brazil. Veterinary Parasitology 153(1-2) 19–23.

Öğdül, H.G. Urban and rural definitions in regional context: a case study on Turkey. European Planning Studies, v.18, n.9, p.1519–1541, 2010.

Okumu F.O., Govella N.J., Moore S.J., Chitnis N., Killeen G.F. (2010) Potential benefits, limitations and target product-profiles of odor-baited mosquito traps for malaria control in Africa. PLoS One 5(7) e11573.

Okwor I., Uzonna J. (2016) Social and Economic Burden of Human Leishmaniasis. The American journal of tropical medicine and hygiene 94(3) 15–0408.

Pan American Health Organization: Leishmaniasis: Epidemiological Report in the Americas: Washington: Pan American Health Organization; 2019. Available at: www.paho.org/leishmaniasis

Pigott D.M., Bhatt S., Golding N., Duda K.A., Battle K.E., Brady O.J., Messina J.P., Balard Y., Bastien P., Pratlong F., Brownstein J.S., Freifeld C.C., Mekaru S.R., Gething P.W., George D.B., Myers M.F., Reithinger R., Hay S.I. (2014) Global distribution maps of the leishmaniases. Elife. 3 e02851.

Poché D.M., Torres-Poché Z., Garlapati R., Clarke T., Poché R.M. (2018) Shortterm movement of Phlebotomus argentipes (Diptera: Psychodidae) in a visceral leishmaniasis-endemic village in Bihar, India. Journal of Vector Ecology. 43(2): 285–92.

Rangel E.F. Shaw J.J. (2018) Brazilian Sand Flies Biology, Taxonomy, Medical Importance and Control. Springer International Publishing.

Reiner R.C. Jr, Stoddard S.T., Forshey B.M., King A.A., Ellis A.M., Lloyd A.L., Long K.C., Rocha C., Vilcarromero S., Astete H., Bazan I., Lenhart A., Vazquez-Prokopec G.M., Paz-Soldan V.A., McCall P.J., Kitron U., Elder J.P., Halsey E.S., Morrison A.C., Kochel T.J., Scott T.W. (2014) Timevarying, serotype-specific force of infection of dengue virus. Proc Natl Acad Sci USA. 111(26) E2694–702.

Reithinger R., Coleman P.G., Alexander B., Vieira E.P., Assis G., Davies C.R. (2004) Are insecticide-impregnated 16 dog collars a feasible alternative to dog culling as a strategy for controlling canine visceral leishmaniasis in brazil? International Journal for Parasitology 34(1) 55–62.

Ribas L.M., Zaher V.L., Shimozako H.J., Massad E. (2013) Estimating the optimal control of zoonotic visceral leishmaniasis by the use of a mathematical model. The Scientific World Journal 1–6.

Roberts G.O, Rosenthal J.F. (2009) Examples of adaptive MCMC. Journal of Computational and Graphical Statistics 18(2) 349–367.

Rock K.S., Quinnell R.J., Medley G.F., Courtenay O. (2016) Progress in the Mathematical Modelling of Visceral Leishmaniasis. In: Basáñez, M.G., Anderson, R.M., (Eds.), Mathematical Models for Neglected Tropical Diseases: Essential Tools for Control and Elimination, Part B, pp. 49–131

Rock K.S., le Rutte E.A., de Vlas S.J., Adams E.R., Medley G.F., Hollingsworth T.D. (2015) Uniting mathematics and biology for control of visceral leishmaniasis. Trends Parasitol. 31:251–9.

Quinnell R.J., Dye C. (1994) An experimental study of the peridomestic distribution of Lyzomyia longipalpis (Diptera, Psychodidae). Bulletin of Entomological Research 84(3): 379–82.

Saul A. (2003) Zooprophylaxis or zoopotentiation: The outcome of introducing animals on vector transmission is highly dependent on the mosquito mortality while searching. Malar J 2: 32.

Schneider M.C., Santos-Burgoa C., Aron J., Munoz B., Ruiz-Velazco S., Uieda W. (1996) Potential force of infection of human rabies transmitted by vampire bats in the Amazonian region of Brazil. Am J Trop Med Hyg. 55(6) 680–4.

Shimozako H.J., Wu J., Massad E. (2017) The preventive control of zoonotic visceral leishmaniasis: Efficacy and economic evaluation. Computational and Mathematical Methods in Medicine 1–21.

Smith T., Maire N., Dietz K., Killeen G.F., Vounatsou P., Molineaux L., Tanner M. (2016) Relationship between the entomologic inoculation rate and the force of infection for Plasmodium falciparum malaria. Am J Trop Med Hyg. 75(2 Suppl) 11–8.

Solano-Gallego Solano-Gallego L., Miro G., Koutinas A., Cardoso L., Pennisi M.G., Ferrer L., Bourdeau P., Oliva G., & Baneth G. (2011) LeishVet guidelines for the practical management of canine leishmaniosis. Parasites & Vectors 4 86.

Sousa-Paula L.C., Silva L.G.D., Sales K.G.D.S., Dantas-Torres F. (2019) Failure of the dog culling strategy in controlling human visceral leishmaniasis in Brazil: A screening coverage issue? PLoS Negl Trop Dis. 13(6) e0007553.

Sundar, S. and Singh, A. (2017). Chemotherapeutics of visceral leishmaniasis: present and future devel-opments. Parasitology, 145(4):481–489.

Teixeira-Neto R.G., da Silva E.S., Nascimento R.A., Belo V.S., de Oliveira C., Pinheiro L.C., Gontijo C.M. (2014) Canine visceral leishmaniasis in an urban setting of Southeastern Brazil: an ecological study involving spatial analysis. Parasit Vectors 7 485.

Thornton, S. J., Wasan, K. M., Piecuch, A., Lynd, L. L. D., and Wasan, E. K. (2010). Barriers to treatment for visceral leishmani-asis in hyperendemic areas: India, bangladesh, nepal, brazil and sudan. Drug Development and Industrial Pharmacy, 36(11):1312–1319.

Trapero-Bertran, M., Mistry, H., Shen, J., and Fox-Rushby, J. 2012. “A Systematic Review and Meta-analysis of Willingness-to-Pay Values: The Case of Malaria Control Interventions.” Health Economics.

Torres-Guerrero, E., Quintanilla-Cedillo, M. R., Ruiz-Esmenjaud, J., and Arenas, R. (2017). Leishmaniasis: a review. F1000Research, 6:750.

Werneck G.L., Costa C.H., Walker A.M., David J.R., Wand M., Maguire J.H. (2007) Multilevel modelling of the incidence of visceral leishmaniasis in Teresina, Brazil. Epidemiol Infect 135(2):195–201.

Werneck, G. L., Costa, C. H. N., de Carvalho, F. A. A., do Socorro Pires e Cruz, M., Maguire, J. H., and Castro, M. C. (2014). Effectiveness of insecticide spraying and culling of dogs on the incidence of leish-mania infantum infection in humans: A cluster randomized trial in teresina, brazil. PLoS Neglected Tropical Diseases, 8(10):e3172.

On the Control of the Leishmaniases & World Health Organization, W. E. C. (2010). Control of the leishmaniases: report of a meeting of the who expert commitee on the control of leishmaniases, Feneva, 22-26 march 2010. In World Health Organ Tech Rep Ser., volume 949.

WHO/Department of Control of Neglected Tropical Diseases (2017) Global leishmaniasis update, 2006–2015: a turning point in leishmaniasis surveillance. Wkly. Epidemiol. Rec. 92, 557–565

Cost effectiveness and strategic planning (WHO-CHOICE). World Health Organization; Available from: http://www.who.int/choice/en/ [accessed 8/2/20].

Williams B.G., Dye C. (1994) Maximum likelihood for parasitologists. Parasitol Today. 10(12) 489–93.

Wilson AL, Courtenay O, Kelly-Hope LA, Scott TW, Takken W, Torr SJ, et al. (2020) The importance of vector control for the control and elimination of vector-borne diseases. PLoS Negl Trop Dis 14(1): e0007831. https://doi.org/10.1371/journal.pntd.0007831

https://data.worldbank.org/indicator/NY.GDP.PCAP.CD?locations=BR

Zahid M.H., Kribs C.M. (2020) Impact of dogs with deltamethrin-impregnated collars on prevalence of visceral leishmaniasis. Infect Dis Model. 5: 235–247.

Zuben A.P.B. von, Donalísio M.R. (2016) Dificuldades na execução das diretrizes do Programa de Vigilância e Controle da Leishmaniose Visceral em grandes municípios brasileiros. Cad. Saude Publica 2016; 32: 1–11.

https://www.who.int/leishmaniasis/resources/who_paho_era6/en/

https://www.paho.org/hq/dmdocuments/2013/Report-Leishmaniasis-PAHO-Eng.pdf?ua=1

