## Supporting Information for "Spatial modelling of sand fly vector’s response to a synthetic sex-aggregation pheromone: impact on the incidence of visceral leishmaniasis in rural and urban settings"

### A1. Spatial distribution and preferences for attractors of sand flies

We have defined an attraction profile as:

$$A^S(d,n,\boldsymbol{p}) = exp(-p_1 d)p_2^S\left(1 - \exp\left(-p_3^S n\right)\right),$$

where $d$ is a distance, $n$ is either a number of pheromone lures (each lure equipped with 10mg of pheromone) or a number of hosts; and $\boldsymbol{p} = \{p_1, p_2^S, p_3^S\}$ are parameters defining the shape of attraction profile for an attractor $S = \{P, H, D, C\}$, where pheromone (P), human host (H), dog (D) or chicken (C) indicates a type of attractor. We made the following assumptions: (i) attraction decreases with a distance from a source with an exponential decay defined by a parameter $p_1$; (ii) there is a saturation effect with increasing amount of pheromone in lures or hosts which is defined by parameter $p_3^S$; (iii) the height of the attraction profile is equal to $p_2^S$. The behaviour of a sand fly responding to a source of attraction in the field depends on the distance required to travel and the strength of stimulus received.
Therefore, we assume that a proportion of sand flies attracted to a household $h$ with $n_S(t)$number of attractors at time $t$ can be calculated by summing the integrals:

$$f_h(t) = \frac{\sum_{S\in\{P,H,D,C\}} \int_0^\infty K(x) A^S(|x^h - x|, n_S(t), \mathbf{p}^S)\, dx}{F(t)},$$

where *K(x)* is the dispersal kernel (details in SI Dispersal of sand flies), and $x^h$ are coordinates of the household *h*.
Normalisation is obtained by summing over all households, i.e. where

$$F(t) = \sum_h \sum_{S\in\{P,H,D,C\}} \int_0^\infty K(x) A^S(|x^h - x|, n_S(t), \mathbf{p}^S)\, dx.$$

We can also calculate a fraction of sand flies attracted to a particular host in a household *h*, for example for dogs:

$$f_h^D(t) = \frac{\int_0^\infty K(x) A^D(|x^h - x|, n_D(t), \mathbf{p}^D)\, dx}{F(t)}.$$

### A2. Dispersal of sand flies

The dispersal of *Lutzomyia longipalpi*s, the important sand fly vector throughout Latin America, was evaluated through the mark-release-recapture method [7]. The relationship between the percentage of recaptured sand flies (Iog scale) has been estimated to be proportional to $-0.16\sqrt{d}$, where $d$ is a distance in meters. Consequently, we have set the dispersal kernel function as:

$$K(d) = K_0 \exp\left(-0.16\sqrt{d}\right),$$

where

$$K_0 = \left(\int_0^\infty exp\left(-0.16\sqrt{x}\right)dx\right)^{-1}$$

Figure S1 shows experimental data [7] and the dispersal kernel *K(d)* as a function of a distance.

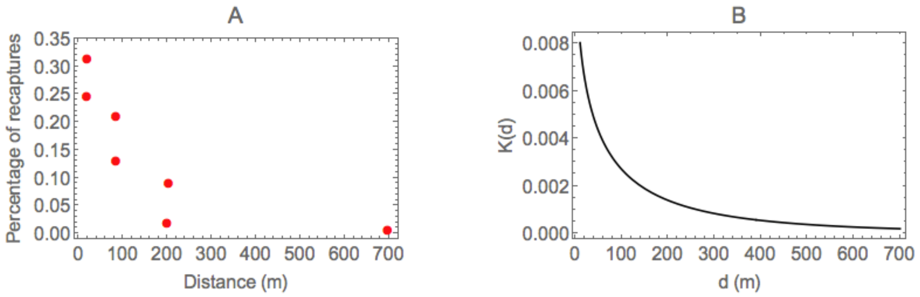


**Figure S1**: Dispersal of sand flies: (A) proportions of re-captured sand flies as a function of distance from the realise point [7]. (B) transmission kernel *K(d)*.

### A3. Fitting attraction profiles to the data

There are nine parameters defining attraction profiles: $\{p_1, p_2^P, p_3^P, p_2^H, p_3^H, p_2^P, p_3^D, p_2^C, p_3^C\}$, where the upper indices are for pheromone (P), human hosts (H), dogs (D) or chickens (C). We set $p_2^P = 1$, i.e. the height of the attraction profile for a maximum amount of pheromone lures to be equal to one.
We have used three sets of available experimental data:
**Experiment A:** Attraction of sand flies to synthetic pheromone: capture experiments [1]. The attraction of individual sand flies to different concentrations of pheromone was measured using a series of choice tests. The experiments aimed to test the dose response to synthetic pheromone relative to chicken-only controls. The quantities of synthetic pheromone tested were 1, 5, 10 or

50 mg and at distances of 5, 10, 15, 20 and 30 meters between test and control CDC light traps [1].

**Experiment B:** Attraction of sand flies to synthetic pheromone: re-capture experiments [20]. These experiments were done using sand flies which were marked with fluorescent powders and released at a specific distance from the test trap. Lures were loaded with 10 mg of synthetic pheromone and the distance was set to 5, 10, 15, 20 and 30 meters between release point and a CDC light trap [20].

**Experiment C:** Host preference experiments [5]. Fieldwork approach was used to investigate the preference of sand flies for dogs, humans and chickens. CDC light traps were set in three domestic locations: one trap in the bedroom of the house, one in the chicken shed and one trap in a dog cage. Numbers of sand flies captured per night in each trap were calculated [5].

In the experiment A, there were two chicken sheds positioned at $x^{test}$ and $x^{control}$ sites. In each shed, there was a synthetic pheromone trap equipped with $\varphi_i$ *(i={contr, test})* amount of synthetic pheromone, where $\varphi_i = 0$ corresponds to the control with no synthetic pheromone present. We set up a virtual experiment within a rectangle around a control and test trap and assumed that sand fly can enter at a random point along the perimeter. The proportion of sand flies attracted to the test trap is calculated as:

$$\alpha_j^A = \frac{\beta(d_{test}, \phi_{test}, x^{test})}{\beta(d_{test}, \phi_{test}, x^{test}) + \beta(d_{contr}, \phi_{contr}, x^{control})},$$

$$\beta(d, \phi, y) = \int_0^d K(x) A(|y - x|, \phi, n_C, \mathbf{p})\, dx/d,$$

where $d_{test}$ is a distance between the test trap and entry point; $d_{control}$ is a distance between a control trap and entry point. As a chicken was placed in each shed to provide a source of host odour, we assume that:

$$A(|y - x|, \phi, n_c, \mathbf{p}) = A^P(|y - x|, \phi, \mathbf{p}) + A^C(|y - x|, 1, \mathbf{p}),$$

i.e. the attraction profile is a sum of attraction profiles for synthetic pheromone and chickens with nC = 1. We run r simulations with randomly positioned entry point along perimeter of the rectangle and set $\alpha^A = \frac{1}{r}\sum_{j=1}^{r} \alpha_j^A$.

In experiment B, sand fly was captured, colour-coded, released and recaptured. We assumed that released sand flies which were not re-captured in the test trap, have dispersed elsewhere. Therefore the proportion of sand flies attracted to the test trap was calculated as

$$\alpha^B = \frac{\int_{\mathcal{R}^2} K(\mathbf{y}) A(|\mathbf{x} - \mathbf{y}|, \phi, n_c, \mathbf{p})\, d\mathbf{y}}{\int_{\mathcal{R}^2} K(\mathbf{y}) A(|\mathbf{x} - \mathbf{y}|, \phi, n_c, \mathbf{p})\, d\mathbf{y} + \int_{\mathcal{R}^2} K(\mathbf{y})\, d\mathbf{y}}$$

In experiment C, the proportions of sand flies attracted to humans, dogs and chickens are calculated as:

$$\alpha_{s,i}^C = \frac{(1/d_s) \int_0^{d_s} K(x) A(|x^s - x|, n_s, \mathbf{p})\, dx}{\sum_{s \in S} (1/d_s) \int_0^{d_s} K(x) A(|x^s - x|, n_s, \mathbf{p})\, dx},$$

where $S = \{H;D;C\}$, $d_S$ is a distance between a trap S and entry point, $n_S$ is a number of individuals of type $S$. We run r simulations with randomly positioned entry point and $\alpha^C = \frac{1}{r}\sum_{j=1}^{r} \alpha_{S,i}^C$

Finally, we assume that the number of sand flies caught in a trap follows a binomial distribution with success probability equal to $\alpha^A$ for the experiment A, $\alpha^B$ for the experiment B, and multinomial distribution with probabilities $\alpha^C$ for the experiment C. Then the likelihood function is given by:

$$\begin{aligned}\mathcal{L}(\boldsymbol{p}) &= \prod_{i=1}^{n_A} Bin(n_i^{test}, n_i^{test}+n_i^{contrl}, \alpha^A) \times \prod_{i=1}^{n_B} Bin(n_i^{recapture}, n_i^{released}, \alpha^B). \\ &\times \prod_{i=1}^{n_C} Multin(n_i^C, n_i^D, n_i^H, n_i^H+n_i^D+n_i^C, \alpha^C) \qquad (9)\end{aligned}$$

We used the MCMC algorithm to estimate the parameters from 309 available data sets. We set a number of entry points to r = 105. We chose uniform priors U[0; 1]. Parameters were updated using an adaptive random walk Metropolis algorithm with proposal distribution given at iteration k [15]:

$$Q_k(x,.) = \begin{cases}\mathcal{N}\left(x,(0.1)^2\mathcal{I}_m/m\right), & \text{if } k \le 2m \\ (1-\xi)\mathcal{N}\left(x,(2.38)^2\Sigma_m/m\right)+\xi\mathcal{N}\left(x,(0.1)^2\mathcal{I}_m/m\right), & \text{otherwise}\end{cases}$$

where $m = 8$ is a number of parameters to estimate, $\Sigma_m$ is an empirical estimate of the covariance matrix at iteration k, and $\xi = 0.05$.

Figure S2 shows the log-likelihood trace plot for the final 1,000 MCMC iterations and posterior distributions of attraction parameters.
Experimental data and fitted proportions are shown in Figure S3. A greater proportion of sand flies would be attracted to a trap with synthetic pheromone relative to a control trap with only a chicken host, and this proportion depends on an amount of lures and showed no dependence on the distance between test and control traps. Fitted attraction profiles indicate that the number of *Lu.longipalpis* attracted to the synthetic pheromone can be increased through addition of more synthetic pheromone, but the effect saturates at about 10 mg per trap (Figure 3S(C)).
Fitted attraction profiles calculated using mean values of parameters from the posterior distribution are shown in Fig.4S.

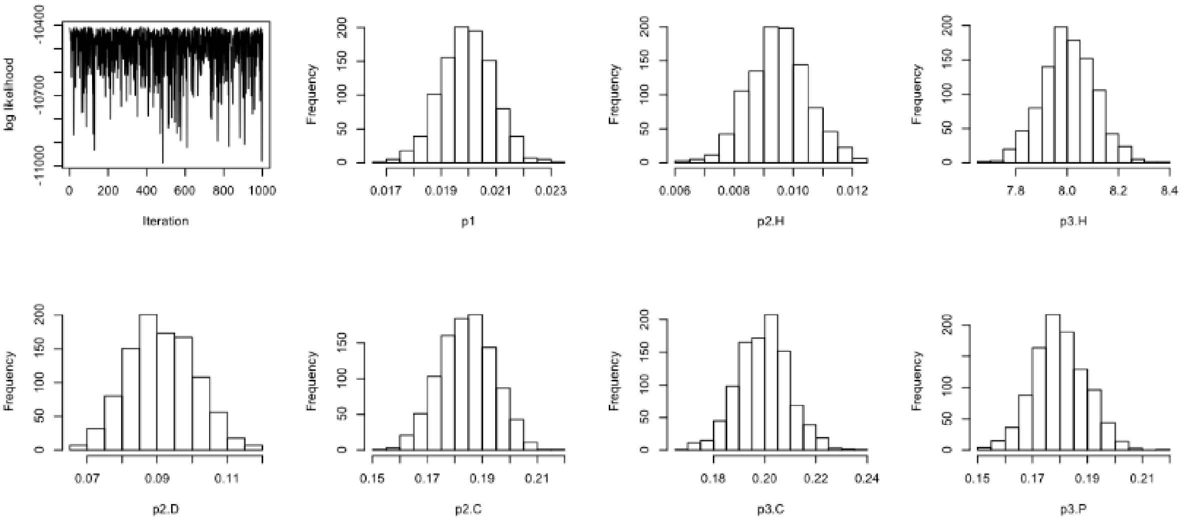


**Figure S2:** Output of the MCMC algorithm when fitting parameters of attraction profiles to the experimental data.

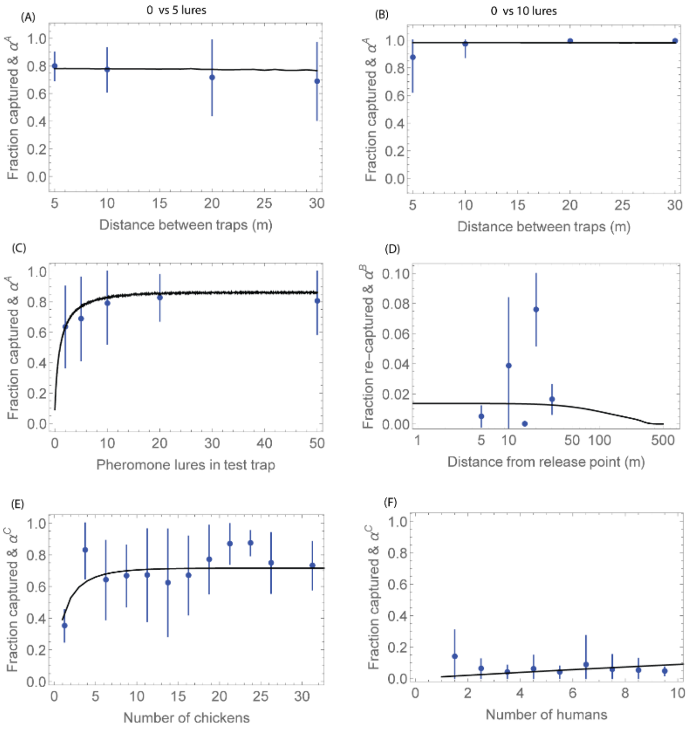


**Figure S3**: Proportions of sand flies attracted to test traps. Measured mean and standard deviation are shown in blue and fitted curves in black: (A)-(D) synthetic pheromone lures; (E) chickens and (F) humans.

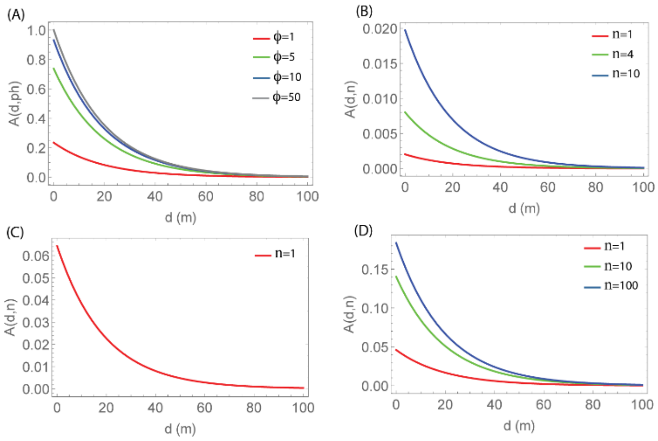


**Figure S4:** Fitted attraction profiles as a function of distance attracted to: (A) synthetic pheromone lures; (B) humans; (C) dogs; and (D) chickens.

### A4. Temporal distribution of sand flies

As a measure of relative sand fly abundance, we used the number of sand flies per trap per day captured by CDC light trap in Boa Vista village, Marajó, Pará state, in Northern Brazil [5]. The seasonality was incorporated as a sinusoidal function of the form:

$$V(t) = b_0\big(1 + b_1 cos(2\pi(t - b_2)/365)\big),$$

where $b_1$ is the amplitude of seasonality, and $b_2$ is a time shift for a peak activity of sand flies. Equation (10) was fitted to the data using the least squares method and gave the following values: $b_0$ = 270, $b_1$ = 0.653576, and $b_2$ = 8. Sand fly trapping data [5] and fitted curve are shown in Fig. 5S(A).

As the absolute sand flies density is unknown and the trapping data only indicates the relative sand flies density pattern, we introduce a scaling parameter . Number of vectors at a household h at time t is given by:

$$V_h(t) = \gamma n V_r(t) f_h \ (t)$$

where n is a number of households.

We have estimated scaling using measured *L. infantum* infection prevalence in dogs on the Marajó [5] (details in SI Estimating the absolute abundance of sand flies). An example of the absolute sand flies abundance for $\gamma = 1.73$ is shown in Fig. 5S(B).

We can also calculate a number of sand flies attracted to a particular host in a household h at time t, for example for dogs:

$$V_h^D(t) = \gamma n V_r(t) f_h^D(t)$$

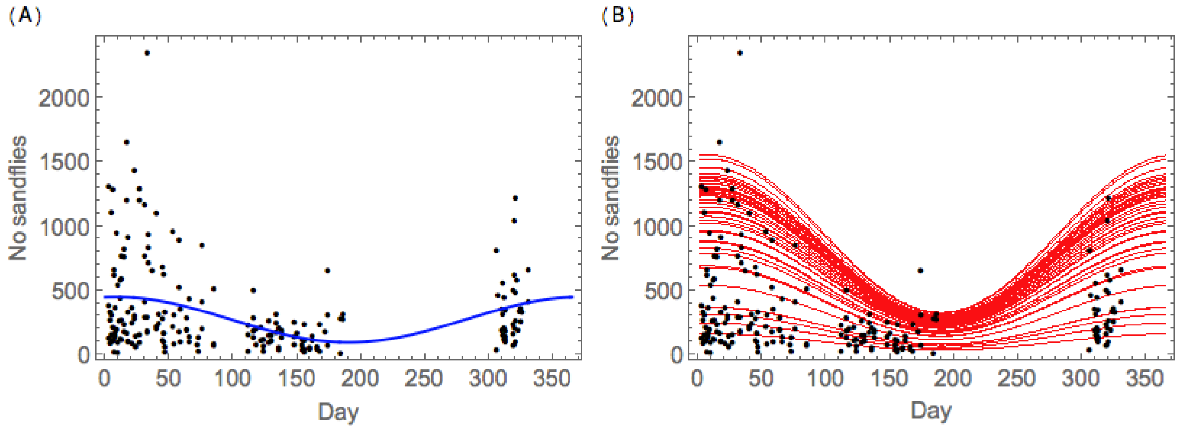


**Figure S5:** Temporal distribution of sand flies. (A) relative sand flies abundance, with fitted equation shown in blue; (B) absolute sand flies abundance, Vh(t), with each red curve corresponding to an individual household. Black dots show sandy trapping data from Boa Vista village [5].

### A5. Spatially targeted interventions

To account for a local impact of synthetic pheromone lures, we apply following steps:

1. We calculate a number of sand flies in each household without pheromone lures $\widehat{V_h(t)}$.
2. For each household with a pheromone lure, we re-distributed sand flies to pheromone lure and hosts according to a ratio between attraction profiles:

$$f_s = \frac{\int_0^\infty K(x) A^s(|x^s - x|, n_s(t), \mathbf{p}^s)}{\sum_{S \in \{P,H,D,C\}} \int_0^\infty K(x) A^S(|x^S - x|, n_S(t), \mathbf{p}^S)}.$$

3. Number of sand flies attracted to a hosts after introduction of synthetic pheromone is equal:

$$V_h^s(t) = f_s \hat{V}_h(t).$$

### A6. Transmission of *L. infantum* infection to dogs

We assume that dog population consist of four types of animals: susceptible, exposed, never infectious, low infectiousness, and high infectiousness.

Each dog in the model has a number of attributes:

i) House of residency. A house where a dog stays over night.

ii) Date of introduction.

iii) Infectiousness status. Each dog at a time of introduction is randomly assigned a status as highly infectiousness, low infectiousness or never infectious. The proportions of never

infectious, low infectiousness, and high infectiousness are $\pi_{never}$, $\pi_{low}$ and $\pi_{high}$, respectively.
iv) Date when dog is infected. We assume that latent period has a Poisson distribution with rate $\nu$.
v) Date when dog becomes infectious (for hight and low infectiousness dogs).
vi) Date dog is removed. All dogs are removed due to a death. We assume that dogs don't show symptoms and therefor are not culled due to being identified as infected. However, we assume that there is a difference in mortality rates: exposed and infectious dogs have mortality rate $\delta_i$, and susceptible dogs have mortality rate $\delta$, with $\delta_i > \delta$.
Deceased dogs are replaced after $R_D$ days on average and newly introduced dogs have a probability of $p_{new}$ of being already infected.
The *L. infantum* transmission model is based on a localised vectorial capacity of the sand fly population to transmit infection between dogs [6, 9]:

$$C_h^D(t) = \frac{V_h^D(t)}{N_h^D(t)} \frac{a_D^2 \exp^{-\mu\tau}}{\mu},$$

where $C_h^D(t)$ is a number of sand flies attracted to dogs at household $h$, $N_h^D(t)$ is a number dogs at household $h$ at time $t$, $a_D$ is a biting rate of female sand fly on dogs, $\mu$ is sand fly mortality rate, and $\tau$ is latent period of in sand flies .
We have modified the force of infection [6, 9] on a dog in a household $h$ at time $t$ so it includes a spatial landscape of infection in dogs:

$$\lambda_h^D(t) = \frac{p_D C_h^D(t)}{N_h^D(t)} \left( p_V^{HI} \sum_{j \in \Omega_{HI}(t)} I_j^{HI}(t) K(d_{h,j}) + p_V^{LI} \sum_{j \in \Omega_{LI}(t)} I_j^{LI}(t) K(d_{h,j}) \right)$$

Here $p_D$ is a probability of an infected sandy transmitting to a dog, $p_V^{HI}$ p and $p_V^{LI}$ p are probabilities of high and low infectiousness dogs transmitting to a sand fly, $\Omega_{HI}(t)$ and $\Omega_{LI}(t)$ are sets of households with high and low infectiousness dogs at time $t$; $I_j^{HI}(t)$ and $I_j^{LI}(t)$ are numbers of high and low infectiousness dogs at household $j$; and $d_{h,j}$ is distance between a household $h$ and a household where dog $j$ resides.

### A6. Number of infectious bites in humans

Number of infectious bites in a house h per human host at time t is calculated as:

$$n_{ib}(t) = \frac{C_h^H(t)}{N_h^H(t)} \left( p_V^{HI} \sum_{j \in \Omega_{HI}(t)} I_j^{HI} K(d_{h,j}) + p_V^{LI} \sum_{j \in \Omega_{LI}(t)} I_j^{LI} K(d_{h,j}) \right)$$

A localised vectorial capacity is equal [6, 9]:

$$C_h^H(t) = \frac{V_h^H(t)}{N_h^H(t)} \frac{a_D a_H \exp^{-\mu\tau}}{\mu},$$

where $a_H$ is biting rate of female sand y on humans.

### A7. Rural and urban settings

The spatial and demographic data for a rural setting came from the fieldwork and survey data from Calderao village, Marajó [5]. The data consisted of GPS coordinates of households, and the number of humans, dogs and chickens resident in the household. Households with missing data for human numbers were assigned a median of five closest neighbours, and number of chickens were sampled from the Marajó dataset. The latitude and longitude coordinates of households were converted to Universal Transverse Mercator zone 22 projection and translated

so that the left bottom corner of a local coordinate system is (0,0). There were 235 households in the village.
The GPS locations and demographic data for the urban setting came from Governador Valadares, Minas Gerais state, Brazil, previously described [19] including data from four urban surveyed blocks. Table S1 shows a comparison of summary statistics between rural and urban setting.
To account for transmission coming from surrounding households, we have fitted rectangles 100m away from each block and assumed that there's an external infection on each household equal to an average infection from within a block scaled by K(d), where d is a distance from a household to the rectangle.

**Table S1.** Household characteristics.

| **Characteristics** | **Rural setting** | **Urban setting** |
|---|---|---|
| Number of households | 235 | 342 |
| Average no of humans per household | 4.67 | 4.19 |
| Average no of dogs per household | 1.09 | 0.8 |
| Average no of chickens per household | 6.8 | 1.43 |

### A8. Sand flies distribution in urban setting

We have compared a number of sand flies caught in Araçatuba city to the number of sand flies caught in rural Marajó.. Distributions of $\log_{10}$ are shown in Figure S6. The mean of the number of sand flies caught in rural setting is 159.5 per household, while the number of sand flies caught in urban settings is 4.6 per household. This gives the ration between means of $r_{urban} = 0.028$. Therefore, for simulations in urban setting we have rescaled seasonality function by $r_{urban}$.

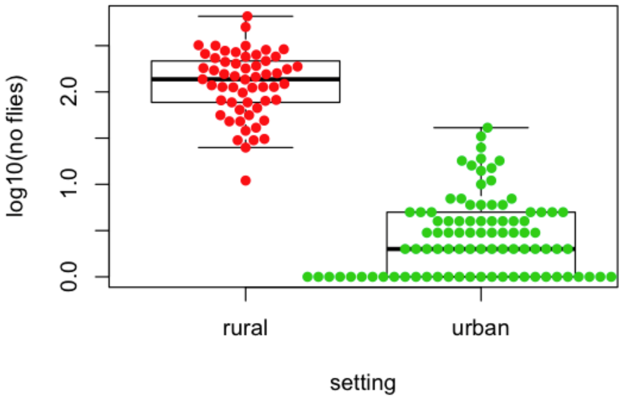


**Figure S6.** Number of trapped sand flies in rural and urban setting.

### A9. Model simulations

The model was simulated using the tau-leaping algorithm with time intervals of one day [8]. At every time t of the simulation (time step  = 1 day), every susceptible dog *i* has a probability of becoming infected:

$$p_i^D(t) = 1 - \exp\left(-\lambda_i^D(t)\right).$$

For each simulation, we randomly chose a single dog as highly infectious and run the sub-model for virus transmission between dogs for ten years till the epidemics reaches endemic

equilibrium. Then we run the model for further ten year with pheromone lures randomly distributed between the households.
The *L. infantum* infection prevalence in dogs at time t is calculated as:

$$pd = 100 \frac{\sum_{j \in \Omega_{HI}(t)} I_j^{HI}(t) + \sum_{j \in \Omega_{LI}(t)} I_j^{LI}(t)}{\sum_{h \in houses} N_h^D(t)}.$$

### A10. Estimating the absolute abundance of sand flies

We have estimated a posterior distribution of parameter using the data on infection prevalence from Marajó and rural setting of the model [5]. We have used approximate Bayesian computation based on a sequential Monte Carlo method [10]. As a distance metrics, we used absolute difference between observed *L. infantum* infection prevalence in dogs and simulated infection prevalence 10 years since the seeded case. The observed infection prevalence in dogs in the rural setting was 49.3% [5].

We used uniform prior distribution $\gamma$~U[0; 100]. The number of desired particles was set to N = 100; we run a single repeat of the model simulation for each proposed parameter set; perturbation kernel was set as a normal distribution with variance calculated using 50 nearest neighbours [10]. We started ABC with tolerances equal to infinity and for later populations the values of thresholds were chosen as the 50th quantile of a posterior distribution from the previous population. We stopped the estimation once the acceptance rate became lower than 5%. Figure S7 (A) shows the posterior distribution for . Model simulations using parameter values from the posterior distribution are shown in Fig. S7 (B).

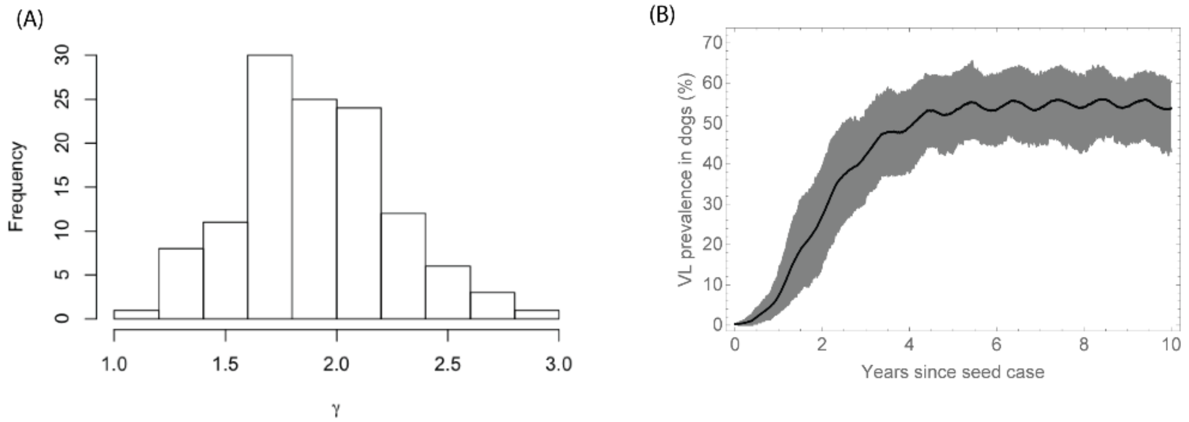


**Figure S7.** Fitting the transmission model to *L. infantum* infection seroprevalence data in dogs: (A) posterior distribution for scaling $\gamma$, (B) simulated infection prevalence in dogs with 100 values of  sampled from a poster distribution.

### A11. Estimating infection exposure in humans

The probability of becoming exposed to *L. infantum* with age was calculated following [17, 2]:

$$P_{sero}(age) = 1 - exp(-P_{inf} \times n_{ib} \times age),$$

where $P_{inf}$ is the probability of being exposed after being bitten by an infected sand fly. Figure S8 shows the fitted probability of humans becoming exposed to *L. infantum* as a function of age. Dots represents prevalences in 1 year age groups from infection exposure measurements in Marajó village  [18].

We have calculated dynamics of infectious bites for each individual in a household pre-control and post-control. We used an approximate age pyramid for Brazil [21] to assign age to all individuals and used probability of being infected after infectious bite to estimate number of new incidences of VL in human population.

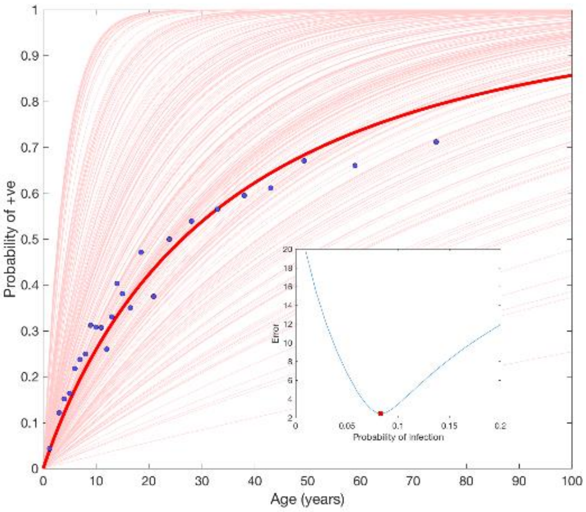


**Figure S8.** Prevalence of *L. infantum* infection exposure in humans against age. Dots represents prevalences in 1 year age groups up to 80 years old [18].

### A12. Control strategies

We initiated simulations with a single infectious dog, run simulations for 10 years until infection prevalence reached endemic equilibrium, and then introduced the synthetic pheromone lure(s) and insecticide in households. We considered the following intervention scenarios: coverage of 50%, 70% and 90% of community households with synthetic pheromone and insecticide; comprising of 1, 5, 10 or 50 lures each containing 10 mg of synthetic pheromone. We also investigated two further scenarios: (i) import of *L. infantum* canine infection via newly introduced dogs, or (ii) import of fully susceptible dogs without infection. We run simulations for further 10 years after the introduction of synthetic pheromone and insecticide.

Figure S9 show simulations of *L. infantum* infection prevalence in dogs for various control options.

Figure S10 shows the relative percentage reduction with no import of infectious dogs. Intervention coverage per settlement (percent of households treated with the synthetic pheromone + insecticide intervention) was set to 50%, 70% and 90%, comprising different number of synthetic pheromone lures 1, 5, 10, or 50, each containing 10 mg of synthetic pheromone.

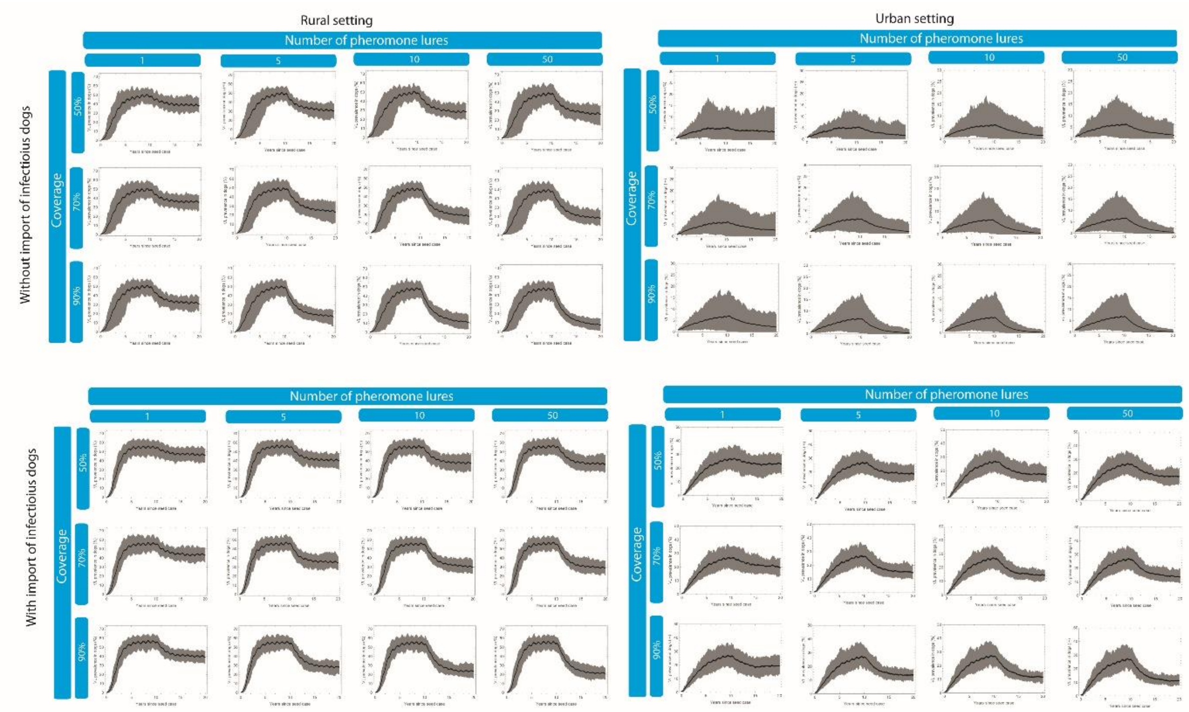


**Figure S9.** The effect of control strategies on *L. infantum* infection prevalence in dogs under different assumptions (with/without import of infectious dogs), settings (rural/urban), combinations of coverage and the amount of synthetic pheromone: mean (black curve) and 95% CI based on 100 outbreak simulations.

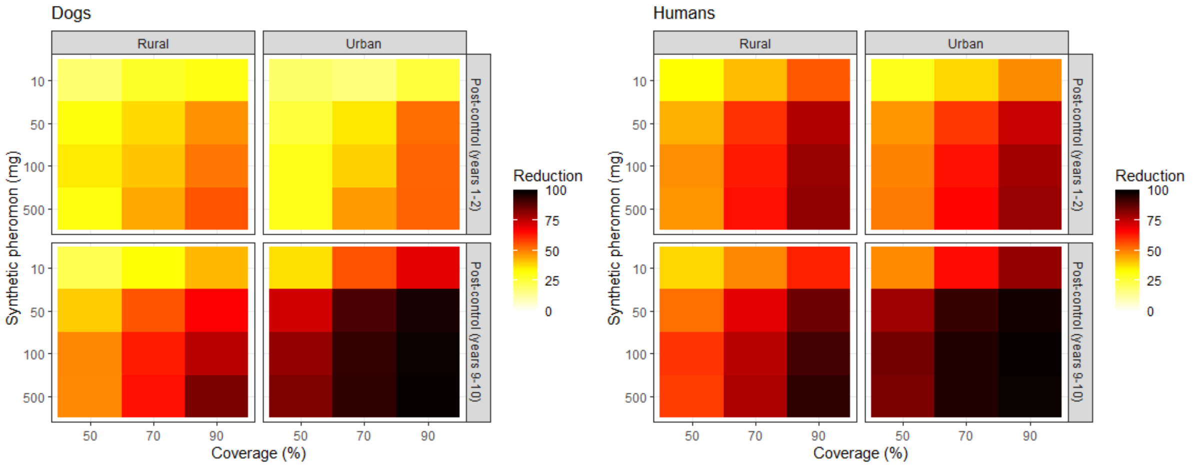


**Figure S10.** Relative percentage reduction with no import of infectious dogs.

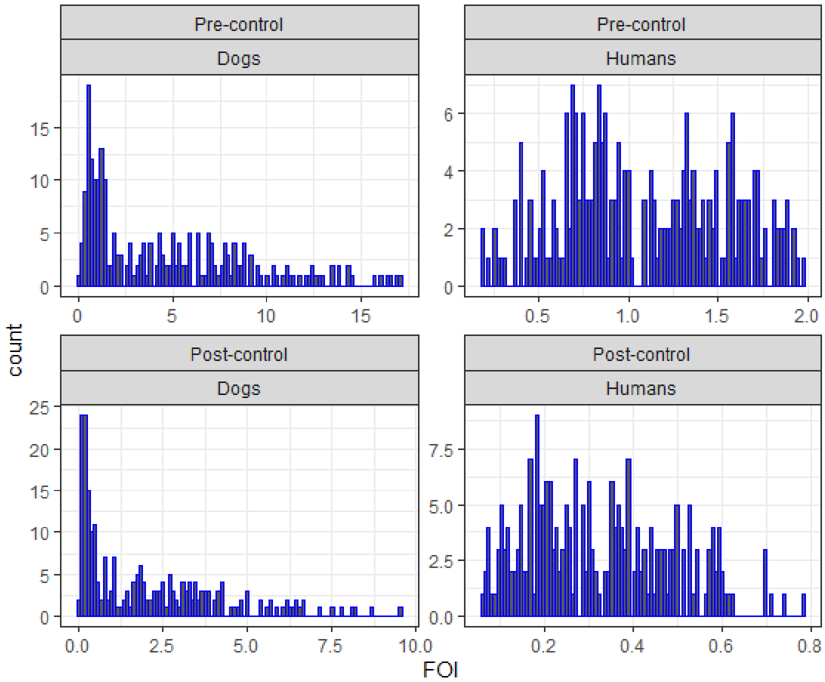


**Figure S11.** Pre-control and post-control distribution of FOI in humans and dogs.

Table S1. List of parameters. TS-this study; d- days.

| Symbol | Description | Value | Source |
|---|---|---|---|
| $b_0$ | Average number of sand flies in a household | 270 | TS |
| $b_1$ | Amplitude of seasonality | 0.653 | TS |
| $b_2$ | Time shift for seasonality | 8 | TS |
| $\mu$ | Sand fly mortality rate ($d^{-1}$) | 0.42 | [6] |
| $\tau$ | Latent period of *L.infantum* in sand flies (days) | 7 | [6] |
| $a_D$ | Biting rate for sand fly on dogs ($d^{-1}$) | 0.333 | [7] |
| $a_H$ | Biting rate for sand fly on humans ($d^{-1}$) | 0.125 | [14] |
| $p_D$ | Probability of an infected sand fly transmitting to a dog | 0.321 | [13] |
| $p_H$ | Probability of an infected sand fly transmitting to a human | 0.08 | TS |
| $p_V^{HI}$ | Probability of high infectiousness dog transmitting to a sand fly | 0.39 | [4] |
| $p_V^{LI}$ | Probability of low infectiousness dog transmitting to a sand fly | 0.017 | [4] |
| $\pi_{high}$ | Proportion of high infectiousness dogs | 0.37 | [3] |
| $\pi_{low}$ | Proportion of low infectiousness dogs | 0.08 | [3, 4] |
| $\pi_{never}$ | Proportion of dogs which never get infected | 0.55 | [4] |
| $p_{new}$ | Probability that newly introduced dogs has been exposed | 0.13 | [11] |
| $\nu$ | Rate of progression of dogs from exposed to infectious ($d^{-1}$) | 0.0055 | [4] |
| $\delta$ | Per capita mortality rate of susceptible dogs ($d^{-1}$) | 0.0011 | [4] |
| $\delta_i$ | Per capita mortality rate of exposed and infectious dogs ($d^{-1}$) | 0.0018 | [16] |
| $R_D$ | Average time for deceased dog to be replaced (*d)* | 121 | [12] |
| $r_{urban}$ | Scaling for a number of sand flies urban to rural setting | 0.028 | TS |

Table S2. Key resources.

| Data set | Source | Application |
|---|---|---|
| Number of recaptured sand flies as a function of distance from source. | [7] | To parametrise vector dispersal kernel. |
| Dose response of sand flies to synthetic pheromone relative to chicken-only controls. | [1] | To parametrise attraction profiles. |
| Re-capture of sand flies released at a specific distance from the test trap. | [20] | To parametrise attraction profiles. |
| Host preference experiments | [5] | To parametrise attraction profiles. |
| Household coordinates and demographic data (Calderao village, Marajó). | [5] | To parametrise spatial model for rural setting. |
| Household coordinates and demographic data (Governador Valadares, Minas Gerais state, Brazil). | [19] | To parametrise spatial model for urban setting. |
| Number of sand flies per trap per day captured by CDC light trap ( Marajó). | [5] | To parametrise vector seasonality shape. |

| | | |
|---|---|---|
| Number of sand flies caught in Araçatuba city | [19] | To estimate scaling of vector abundance between rural and urban setting |
| Prevalence of *L. infantum* in dogs (Marajó). | [5] | To estimate absolute abundance of sand flies in rural setting. |
| Prevalence of *L. infantum* infection exposure in humans against age (Marajó). | [18] | To parametrise VL transmission to humans. |
| Population pyramid for Brazil | [21] | To parametrise VL transmission to humans. |
| Gross domestic product (GDP) for Brazil | [22] | To estimate Willingness to Pay. |

References

[1] Bell, M. J., Sedda, L., Gonzalez, M. A., de Souza, C. F., Dilger, E., Brazil, R. P., Courtenay, O., and Hamilton, J. G. C. (2018). Attraction of *lutzomyia longipalpis* to synthetic sex-aggregation pheromone: Effect of release rate and proximity of adjacent pheromone sources. PLOS Neglected Tropical Diseases, 12(12):e0007007..

[2] O COURTENAY, DW MACDONALD, R LAINSON, JJ SHAW, and C DYE. EPIDEMIOLOGY OF CANINE LEISHMANIASIS - A COMPARATIVE SEROLOGICAL STUDY OF DOGS AND FOXES IN AMAZON BRAZIL. PARASITOLOGY, 109(3):273{279, 1994. doi:{10.1017/ S0031182000078306}.

[3] Orin Courtenay, Connor Carson, Leo Calvo-Bado, Lourdes M. Garcez, and Rupert J. Quinnell. Heterogeneities in leishmania infantum infection: Using skin parasite burdens to identify highly infectious dogs. PLoS Neglected Tropical Diseases, 8(1):e2583, jan 2014. doi:10.1371/journal.
pntd.0002583.

[4] Orin Courtenay, Rupert J. Quinnell, Lourdes M. Garcez, Je_rey J. Shaw, and Christopher Dye. Infectiousness in a cohort of brazilian dogs: Whyculling fails to control visceral leishmaniasis in areas of high transmission. The Journal of Infectious Diseases, 186(9):1314{1320, nov 2002. doi:10. 1086/344312.

[5] Erin Dilger. The effects of host-vector relationships and density dependence on the epidemiology of visceral leishmaniasis. PhD thesis, University of Warwick, 2013.

[6] Christopher Dye. The logic of visceral leishmaniasis control. The American Journal of Tropical Medicine and Hygiene, 55(2):125{130, aug 1996. URL: https://doi.org/10.4269/ajtmh.1996.55.125, doi:10.4269/ajtmh.1996.55.125.

[7] Christopher Dye, Clive R. Davies, and Ralph Lainson. Communication among phlebotomine sandies: a field study of domesticated *Lutzomyia longipalpis* populations in amazonian brazil. Animal Behaviour, 42(2):183{
192, aug 1991. doi:10.1016/s0003-3472(05)80549-4.

[8] Daniel T. Gillespie. Approximate accelerated stochastic simulation of chemicallyreacting systems. The Journal of Chemical Physics, 115(4):1716-1733, jul 2001. URL: https://doi.org/10.1063/1.1378322, doi:10.1063/1.1378322.

[9] Sonia A. Gomez, Lloyd A. C. Chapman, Erin Dilger, Orin Courtenay, and Albert Picado. Estimating the efficacy of community-wide use of systemic insecticides in dogs to control zoonotic visceral leishmaniasis: A modelling study in a brazilian scenario. PLOS Neglected Tropical Diseases, 12(9):e0006797, sep 2018. doi:10.1371/journal.pntd.0006797.

[10] Amanda Minter and Renata Retkute (2019). Approximate bayesian computation for infectious disease modelling. Epidemics 29, 100368 doi: 10.1016/j.epidem.2019.100368.

[11] Edson Duarte Moreira, Verena Maria Mendes de Souza, Meera Sreenivasan, Eliane G_oes Nascimento, and Lain Pontes de Carvalho. Assessment of an optimized dog-culling program

in the dynamics of canine leishmania transmission. Veterinary Parasitology, 122(4):245{252, aug 2004. doi: 10.1016/j.vetpar.2004.05.019.
[12] C_aris Maroni Nunes, Val_eria Mar_cal F_elix de Lima, Henrique Borges de Paula, Silvia Helena Venturoli Perri, Andr_ea Maria de Andrade, Francisca Elda Ferreira Dias, and Marcelo Nascimento Burattini. Dog culling and replacement in an area endemic for visceral leishmaniasis in brazil. Veterinary Parasitology, 153(1-2):19-23, May 2008. URL: https://doi.org/10.1016/j.vetpar.2008.01.005, doi:10.1016/j.vetpar.2008.01.005.
[13] Richard Reithinger, Paul G Coleman, Bruce Alexander, Edvar Paula Vieira, Geraldo Assis, and Clive R Davies. Are insecticide-impregnated dog collars a feasible alternative to dog culling as a strategy for controlling canine visceral leishmaniasis in brazil? International Journal for Parasitology, 34(1):55{62, jan 2004. doi:10.1016/j.ijpara.2003.09.006.
[14] Laila Massad Ribas, Vera Lucia Zaher, Helio Junji Shimozako, and EduardoMassad. Estimating the optimal control of zoonotic visceral leishmaniasis by the use of a mathematical model. The Scienti_c World Journal, 2013:1-6, 2013. URL: https://doi.org/10.1155/2013/810380, doi:10.1155/ 2013/810380.
[15] Gareth O. Roberts and Jeffrey S. Rosenthal. Examples of adaptive MCMC. Journal of Computational and Graphical Statistics, 18(2):349-367, jan 2009. URL: https://doi.org/10.1198/jcgs.2009.06134, doi:10.1198/jcgs.2009.06134.
[16] Helio Junji Shimozako, JianhongWu, and Eduardo Massad. The preventive control of zoonotic visceral leishmaniasis: E_cacy and economic evaluation. Computational and Mathematical Methods in Medicine, 2017:1{21, 2017. doi:10.1155/2017/4797051.
[17] BG WILLIAMS and C DYE. MAXIMUM-LIKELIHOOD FOR PARASITOLOGISTS. PARASITOLOGY TODAY, 10(12):489{493, 1994. doi: {10.1016/0169-4758(94)90163-5}.
[18] O. Courtenay & RJ Quinnell, unpublished data.
[19] Gonçalves et al., in review.
[20] González MA, Bell M, Souza CF, Freitas RM, Brazil RP, Courtenay O, Hamilton JGC (2020). Synthetic sex-aggregation pheromone of Lutzomyia longipalpis, the South American sand fly vector of Leishmania infantum, attracts males and females over long-distance. PLoSNTDs In Press.
[21] https://www.populationpyramid.net/brazil/2019/
[22] https://data.worldbank.org/indicator/NY.GDP.PCAP.CD?locations=BR
